# Treating earlier, spending less: cost-effectiveness and budget impact of immediate versus delayed antiretroviral therapy for HIV in Japan

**DOI:** 10.64898/2026.08.23.26361165

**Authors:** Toshibumi Taniguchi, Mayumi Imahashi, Daisuke Sato, Tatsuya Noda

## Abstract

**Background.:** In Japan, lifelong antiretroviral therapy (ART) is funded through the physical disability (immune dysfunction) certification pathway, which requires two laboratory assessments four weeks or more apart. This statutory pathway, rather than clinical need, contributes to a median diagnosis-to-ART interval of about 42 days. We evaluated relaxing or reforming it to permit immediate ART.

**Methods.:** We developed a stochastic individual-based microsimulation of HIV in Japan, linked to a payer-perspective cost-effectiveness analysis over a 40-year horizon after a 20- year burn-in, calibrated to national surveillance and cascade data. We compared immediate ART with one-month (primary) and two-month (secondary) delays. Costs and quality-adjusted life-years (QALYs) were discounted at 2% per year; uncertainty was assessed across 200 seeds and by probabilistic and one-way sensitivity analyses.

**Findings.:** Against the one-month delay, immediate ART averted 2,991 infections and 1,761 deaths among people with HIV over 40 years, gained 9,071 QALYs, and reduced discounted costs by ¥54·4 billion (net monetary benefit ¥99·7 billion). The two-month comparison saved ¥82·5 billion (4,535 infections, 2,688 deaths averted). Immediate ART was dominant at the base case and in all 1,000 probabilistic sensitivity-analysis iterations; cumulative savings offset the early investment within 11 to 12 years, and sensitivity analyses altered only its magnitude.

**Interpretation.:** Permitting immediate ART by reforming the certification pathway was projected to reduce HIV incidence, improve population health, and save public-payer costs within the second decade, supporting consideration of statutory reform.

**Funding.:** Health and Labour Sciences Research Grant, Ministry of Health, Labour and Welfare of Japan (21HB1003, 23HB1001, 26HB1001).

**Research in context:** 

**Evidence before this study.:** We searched PubMed and Web of Science from database inception to March 31, 2026, without language restrictions, for studies of rapid antiretroviral therapy (ART) initiation, combining the terms "rapid ART", "same-day ART", "early ART initiation", "test and treat", "HIV", and "cost-effectiveness", and we reviewed the World Health Organization (WHO) 2017 and 2021 HIV guidelines together with successive AIDS Surveillance Committee of Japan and Ministry of Health, Labour and Welfare reports.

Randomised trials in sub-Saharan Africa and large implementation programmes, most notably the San Francisco RAPID initiative, have consistently shown that starting ART within one week of diagnosis shortens the time to viral suppression, improves linkage to and retention in care, and reduces onward transmission without increasing early adverse events, and by 2023 99 countries had adopted rapid ART in line with the WHO recommendation. Almost all of this evidence, however, comes from high-prevalence or generalised epidemics; we identified no transmission-dynamic or economic evaluation of rapid ART in a low-prevalence, concentrated epidemic with an already strong treatment cascade such as Japan. Above all, to our knowledge no study had modelled the structural barrier that, rather than any clinical consideration, governs the timing of ART in Japan, namely the statutory immune-dysfunction disability certification required to access subsidised treatment, and earlier Japanese HIV modelling was compartmental and did not represent this pathway.

**Added value of this study.:** To our knowledge, this is the first study to represent the Japanese immune-dysfunction certification pathway explicitly and to quantify, over a 40-year horizon, the epidemiological, clinical, and economic consequences of relaxing it to permit immediate ART. Using an individual-based dynamic transmission model calibrated to the contemporary national epidemic and approximating the observed median diagnosis-to-ART interval, we found that, under the modelled assumptions, immediate ART was dominant: it was projected to avert HIV infections and deaths, improve population health, and lower public-payer costs, supported, for the primary comparison, by dominance in 199 of 200 paired per-seed replicates, a positive net monetary benefit in all 200, and dominance in all 1,000 probabilistic sensitivity-analysis iterations; no prespecified one-way health-economic parameter variation reversed dominance. By showing that earlier treatment is simultaneously more effective and cost-saving even where downstream retention and viral suppression are already high, the study adds a setting that has been largely absent from the global rapid ART evidence base.

**Implications of all the available evidence.:** Taken together with the existing trial and implementation evidence, our findings indicate that relaxing or reforming the statutory certification pathway (allowing ART at diagnosis irrespective of CD4 count and HIV-RNA) could reduce HIV incidence, prevent deaths among people with HIV, and save public-payer resources in Japan, while bringing national practice into line with the WHO recommended standard. Because the barrier is regulatory rather than clinical, policy reform is a potentially actionable route, warranting consideration and further evaluation. More broadly, the results suggest that, even in low-prevalence concentrated epidemics with high treatment coverage, removing administrative delays to ART initiation may deliver population-level health and fiscal benefits.

## Introduction

Human immunodeficiency virus (HIV) infection remains a persistent public health challenge. Antiretroviral therapy (ART) has transformed HIV into a manageable chronic condition and, when initiated promptly with sustained adherence, renders the virus sexually non-transmissible.^1–3^ Since 2017 the WHO has recommended that ART be initiated within one week of confirmed HIV diagnosis for people who are clinically ready, with same-day initiation where appropriate, a policy known as rapid ART.^4,5^ Randomised trials in sub- Saharan Africa^6,7^ and implementation studies in high-income metropolitan settings such as the San Francisco RAPID programme^8,9^ have consistently shown that rapid ART shortens time to viral suppression, improves linkage to care, and reduces onward transmission without increasing early adverse events.^10^ More than ninety countries have adopted treat-all policies, and one-week ART initiation is increasingly standard practice.^5^

Japan is a low-prevalence country with a concentrated HIV epidemic in which most transmission occurs among men who have sex with men (MSM).^11^ The Japanese care cascade performs strongly on retention and suppression: national database analyses document on-treatment retention above 95% and viral suppression above 99% among people with HIV retained in care,^12^ and contemporary single-centre data show a standardised mortality ratio approaching that of the general male population.^13^ However, the first arm of the 95-95-95 target remains challenging: approximately 89·3% of people with HIV in Japan were aware of their status at the end of 2022, still below the 95% target.^14^

In Japan, the interval between diagnosis and ART initiation is longer than the international standard, largely for structural rather than clinical reasons. Because contemporary regimens cost far more than the standard 30% co-payment under universal health insurance would reasonably cover, most people with HIV access ART through the Medical Care Subsidy benefit, which requires a physical disability certificate documenting HIV-related immune dysfunction. Except for individuals presenting with AIDS, certification requires two sets of laboratory measurements at least four weeks apart satisfying specified thresholds (CD4 of 500 cells/microlitre or above with HIV-RNA of 5,000 copies/mL or above, or CD4 below 500 with HIV-RNA below 5,000). In a recent single-centre study at a Designated HIV/AIDS Core Hospital, conducted within the MHLW research programme, the median interval from first visit to ART initiation was 42 days, approximately 1·6% of previously untreated patients could not begin ART because they did not meet the certification criteria, and approximately 19·4% of patients already on ART at their first visit had not obtained the disability certificate.^15^ These observations pose a specifically Japanese policy question: would statutory reform of the certification pathway yield meaningful health and economic benefits in a low-prevalence setting with a strong downstream cascade?

The international evidence for rapid ART comes largely from high-prevalence sub-Saharan African and United States metropolitan settings, neither of which captures the combination of low prevalence, concentrated MSM epidemic, high downstream retention and suppression, and statutory certification pathway that characterises Japan. Previous evaluations of HIV strategies in Japan used compartmental projections^16^ rather than integrated individual-level assessments that jointly simulate testing, certification, ART initiation, retention, and long-term outcomes. We therefore developed an agent-based microsimulation of HIV transmission, progression, and care in Japan, and used it to evaluate the 40-year clinical, epidemiological, and economic impact of permitting immediate ART through relaxation or reform of the certification pathway, with the four-week requirement reflected in the primary comparison.

## Methods

### Study design and overview

We developed a stochastic individual-based (agent-based) dynamic transmission microsimulation of HIV acquisition, diagnosis, and care in Japan, linked to a cost-effectiveness analysis from the healthcare payer perspective. The model represents individuals in two sexual risk populations, MSM and a high-risk heterosexual population that includes a small high-activity core, and simulates monthly HIV transmission, testing and diagnosis, CD4 and viral load trajectories, ART initiation and viral suppression, pre-exposure prophylaxis (PrEP), background and HIV-related mortality, and demographic turnover including migration. The model was run as a 20-year burn-in (calendar years 2004 to 2023) to reach a calibrated contemporary epidemic state, followed by a 40-year analytic horizon (2024 to 2063), chosen to capture the slowly accruing transmission, mortality, and cost consequences of ART timing. The analytic horizon used costs in Japanese yen and discounted both costs and quality-adjusted life-years (QALYs) at 2% per year, consistent with Japanese guidance.^17^ Reporting followed CHEERS 2022,^18^ the ODD protocol,^19^ and ISPOR-SMDM good research practices for dynamic transmission modelling.^20^

### Policy context and definition of scenarios

Certification of immune dysfunction under the physical disability framework provides access to substantial public subsidy of medical costs, which in practice governs when many people with HIV can begin lifelong ART. The certification criteria are described in the Introduction. These requirements impose a structural delay between entry into care and ART initiation, because patients must accrue qualifying laboratory values before certification and subsidised treatment.

We compared three ART-initiation strategies that differed in the certification policy governing both ART eligibility and the time to ART initiation: immediate ART (0 month), in which ART begins at entry into care as would be possible if the disability-certification pathway were relaxed or reformed (for example, by removing the repeat-assessment, four-week, and CD4/HIV-RNA criteria requirements, or by issuing the disability certificate and medical-care subsidy at diagnosis); a one-month delay; and a two-month delay. Because the four-week interval is the statutory minimum, the one-month delay represents current practice operating as efficiently as the law permits; the 0-month versus 1-month comparison is therefore the principal analysis and yields a conservative, lower-bound estimate. Single-centre retrospective data indicate a median interval from entry into care to ART of 42 days;^15^ because the model advances in monthly steps, the one-month and two-month delays bracket this observed value (modelled diagnosis-to-ART medians of about 30 and 61 days) rather than interpolating a single 1·5-month delay. The two-month delay serves as a secondary, upper-bound comparator reflecting practice and regional variation. All other inputs were held identical across scenarios, so that between-scenario differences are attributable solely to the certification policy, that is, the joint effect of removing the eligibility criteria and the associated waiting interval. Common random numbers were used across scenarios within each replicate. The current certification pathway and the three modelled ART-initiation strategies are summarised in Figure 1.

**Figure 1.**
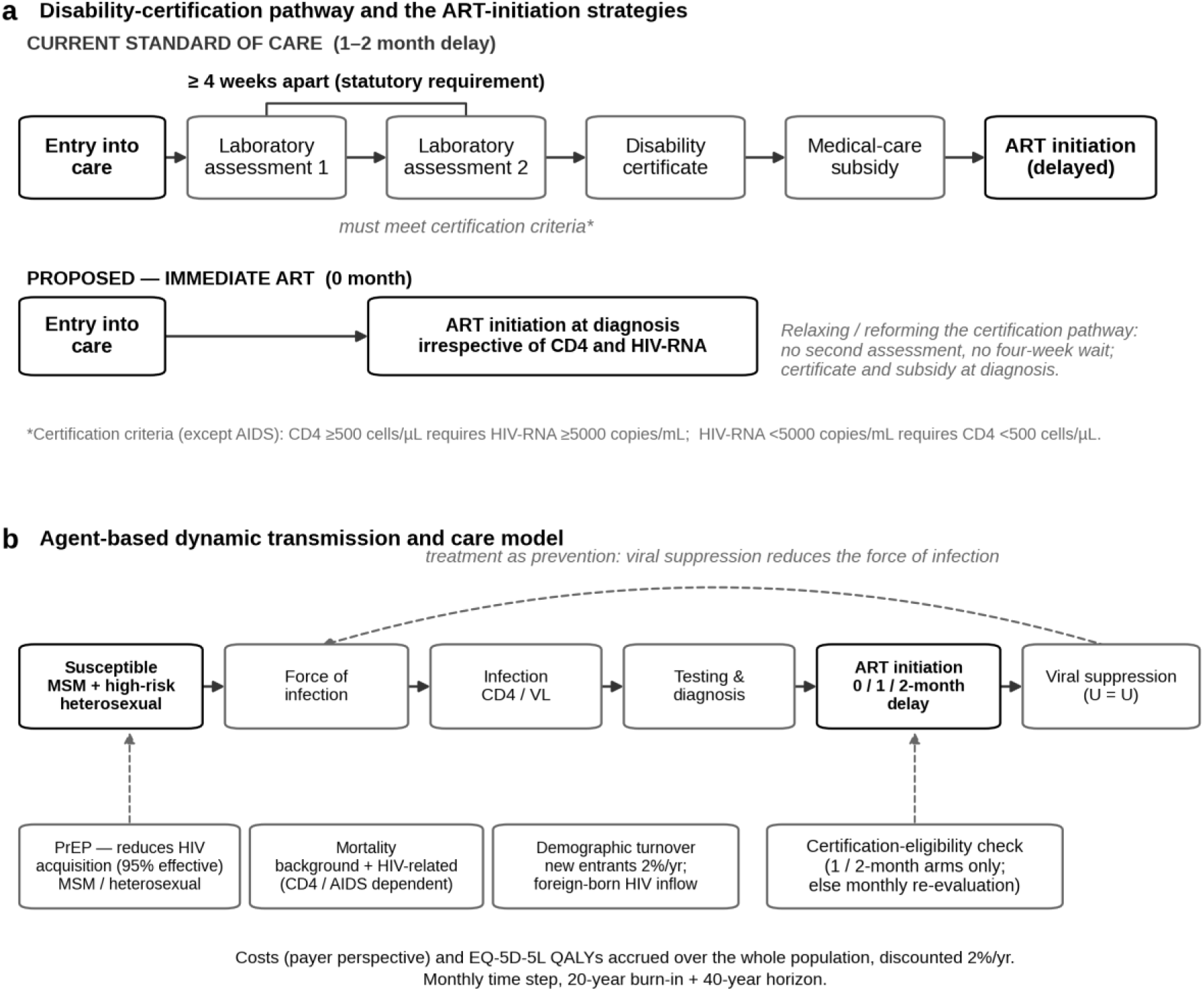
Schematic of the disability-certification pathway and the modelled ART- initiation strategies. Panel a contrasts the current standard of care with the proposed intervention. Under the current pathway, ART is reached only after two laboratory assessments at least four weeks apart that satisfy the immune-dysfunction certification criteria (the CD4 and HIV-RNA threshold combinations), followed by issue of the disability certificate and the medical-care subsidy, producing a one-to-two-month delay. Immediate ART (0 month) starts ART at diagnosis irrespective of CD4 count and HIV-RNA, achieved by relaxing or reforming the certification pathway (for example, removing the repeat- assessment and four-week requirements, or issuing the certificate and subsidy at diagnosis). Panel b shows the agent-based model structure: risk populations (including the high-activity heterosexual core), the force of infection, infection and CD4/viral-load progression, testing and diagnosis, ART initiation under the 0/1/2-month delay, and viral suppression (undetectable equals untransmittable), with treatment as prevention feeding back to reduce the force of infection. Background processes (pre-exposure prophylaxis, which lowers HIV acquisition among susceptibles; mortality; and demographic turnover) and the certification- eligibility check that gates ART initiation in the one-month and two-month arms are also shown. Conceptual diagram; no data.

### Model structure

The simulated population comprised an MSM population and a high-risk heterosexual population, the latter containing a small high-activity core representing concentrated heterosexual transmission (commercial-sex and high-partner-turnover networks). Susceptible individuals acquired HIV through a force of infection that depended on partner contact rates, the viral load distribution of infectious partners, and per-contact transmission probabilities stratified by viral load; per-contact probabilities were anchored on published meta- analyses,^21,22^ with effectively zero transmission at suppressed viral load consistent with PARTNER and Opposites Attract.^1,2^ Diagnosed individuals entered care and initiated ART according to the scenario-specific delay, achieved viral suppression over time, and experienced CD4 recovery; suppression reduced onward transmission consistent with treatment as prevention. Disease progression in the absence of ART followed a five-stage CD4 trajectory with individual stochastic variation, informed by natural-history data.

Mortality combined age- and sex-specific general-population background rates with HIV-related excess mortality represented as relative risks (standardised mortality ratios relative to the general population) jointly stratified by four CD4 strata and four care states (untreated, suppressed on ART, unsuppressed on ART, and disengaged from care); the schedule was anchored to treatment-era MSM cohorts (the Antiretroviral Therapy Cohort Collaboration, Lodwick and colleagues, and COHERE) and checked against a recent Japanese cohort, with values, derivations, and sources in Supplementary Table S1. Demographic turnover included new entrants at an annualised rate of 2·0% and an exogenous foreign-born inflow of people with HIV, with separate rates for the burn-in and main analytic periods. The complete specification is provided as an ODD protocol in Supplementary Material section 1.

### Testing, diagnosis, and ART initiation

Monthly individual testing probabilities were stratified by HIV status, CD4 category, behavioural group, age, and PrEP status, with PrEP users tested quarterly per Japanese guidance.^23^ A national testing-capacity constraint scaled from 14,500 to 18,000 tests per month, reflecting planned national expansion. During the burn-in, testing capacity was anchored to documented national testing volumes, ART initiation followed the historical CD4 eligibility thresholds, and the force of infection carried year-specific calibration factors (Supplementary Material section 2). In the immediate-ART scenario, every diagnosed individual started ART in the month of diagnosis irrespective of CD4 count or HIV-RNA; that is, both the statutory certification criteria and the associated waiting interval were removed. In the comparator scenarios representing current practice (one-month and two-month delays), the certification criteria were applied: individuals meeting the criteria (CD4 below 500 cells/microlitre, or CD4 of 500 or above with HIV-RNA of 5,000 copies/mL or above) were assigned a time to ART drawn from a distribution with mean equal to the scenario target (1 or 2 months) and standard deviation 0·5 months (the eligibility-to-ART interval), whereas individuals not meeting the criteria remained in routine follow-up with monthly re-evaluation until the criteria were met. The model therefore tracks both the eligibility-to-ART and diagnosis-to-ART intervals (Results). The statutory two-assessment, four-week requirement was thus approximated by a single eligibility check at diagnosis, a scenario-specific minimum delay, and monthly re-evaluation of initially ineligible individuals, rather than by explicit simulation of two separate laboratory measurements. Pre-ART and on-ART monthly dropout were applied, informed by Japanese retention data.^12^

### Pre-exposure prophylaxis

Oral PrEP, approved in Japan in 2024 but not publicly funded, was modelled with gradual uptake over ten years to a maximum coverage of 2·0% among MSM and 0·5% among heterosexual individuals, with 95% effectiveness under high adherence^24^ and monthly discontinuation of 0·5%.

### Costs and health-related quality of life

Costs took the public healthcare payer perspective, chosen because the policy change concerns eligibility for publicly subsidised ART and its payer-expenditure consequences. Annual on-ART costs were age-stratified from the publication-cleared National Database of Health Insurance Claims (NDB) aggregate of HIV care costs for fiscal years 2013 to 2020, prepared as part of the MHLW project, consistent with the published NDB cascade update.^12^ Off-ART costs for diagnosed people with HIV were modelled as 45% of on-ART costs, and as 30% in the budget-impact analysis to disregard the diagnosis-to-ART transient cost surge. Health-state utilities were represented as EQ-5D-5L values: Japanese general-population norms by age band for HIV-negative person-time,^25^ and CD4-stratified values for people with HIV on suppressive ART. Because the intervention changes who becomes infected, QALYs were accrued over the whole simulated population so that the benefit of averted infections (person-time lived HIV-negative at higher utility) was captured; restricting QALYs to people with HIV would omit this prevention benefit. Full cost and utility values, distributions, and ranges are given in Supplementary Table S1. Costs are expressed in Japanese yen at the prices of the NDB source period; no adjustment to a single common price year was applied. ^26^ Implementation costs of certification reform were assumed to be negligible because the change is administrative.

### Health-economic analysis

Incremental cost-effectiveness was summarised using the incremental cost-effectiveness ratio (ICER) and the net monetary benefit (NMB) at a willingness-to-pay threshold of ¥5 million per QALY, with NMB also reported across thresholds from 0 to ¥20 million per QALY. A strategy that was both less costly and more effective was classified as dominant. Incremental QALYs were estimated by two consistent methods, of which the prevention decomposition was prespecified as the base case reported in the Abstract, Table 1, and the net monetary benefit, with the whole-population direct estimate reported for comparison. The direct estimate is the single difference of total discounted QALYs from the seed-averaged outputs, requiring no additional decomposition assumptions but forming a small difference of large quantities; the prevention decomposition multiplies averted infections by the discounted QALY loss per infection, plus a treatment-timing component among individuals infected in both strategies, giving a low-variance estimate with confidence intervals. The counterfactual HIV-negative survival weighting was anchored to the whole-population direct estimate, yielding a discounted loss of approximately 2·8 QALYs per averted infection, consistent with near-normal survival under modern ART; the dependence of the health-gain magnitude on this weighting was examined in a one-way sensitivity analysis (Supplementary Material section 4). Incremental costs were computed from cumulative discounted care costs, and the break-even year (the year at which cumulative savings offset the earlier costs of immediate ART, within the modelled payer-cost framework and excluding implementation costs) was determined. The budget-impact analysis projected the cumulative expenditure difference over the full 40-year horizon as a long-term cumulative expenditure projection, discounted at 2% per year; because conventional budget-impact analysis emphasises short-term undiscounted cash flows,^27^ the cumulative undiscounted incremental expenditure over the first five years is also reported.

**Table 1.** Main epidemiological and economic outcomes over the 40-year analytic horizon (hiv_rapid_art_model; N = 200 seeds).

| Outcome | Immediate ART (0 month) | 1-month delay | 2-month delay |
| --- | --- | --- | --- |
| **Epidemiological outcomes (40-year cumulative)** |  |  |  |
| New HIV infections | 13,263 | 16,254 | 17,797 |
| Deaths among people with HIV | 32,576 | 34,337 | 35,263 |
| ART initiations | 25,662 | 27,843 | 29,030 |
| New HIV diagnoses | 15,657 | 17,864 | 18,997 |
| Diagnosis-to-ART, median days (IQR) | 0 (0 to 0) | 30 (30 to 61) | 66 (61 to 91) |
| Diagnosis-to-ART, mean days | ~1 | 120 | 154 |
| Eligibility-to-ART, median days (IQR) | 0 (0 to 0) | 30 (30 to 61) | 61 (61 to 91) |
| Eligibility-to-ART, mean days | 6 | 48 | 76 |
| **Calibration at 2023 (modelled; 95% CI across seeds)** |  |  |  |
| Annual notifications | 911 (823 to 1,000) | shared burn-in |  |
| People on ART | 29,931 (29,042 to 30,820) |  |  |
| Undiagnosed fraction (%) | 11·2 (10 to 12) |  |  |
| **Health-economic outcomes (CEA, discounted)** |  |  |  |
| Total discounted QALYs | 64,197,886 | 64,189,497 | 64,182,715 |
| Total discounted cost (¥ billion) | 2,216·50 | 2,270·88 | 2,299·05 |
| **Incremental outcomes (immediate ART vs delayed)** |  |  |  |
| Infections averted (mean; 95% CI) | reference | 2,991 (2,871 to 3,112) | 4,535 (4,411 to 4,658) |
| Deaths among people with HIV averted (mean; 95% CI) | reference | 1,761 (1,686 to 1,837) | 2,688 (2,610 to 2,766) |
| Incremental QALYs, prevention decomposition, base case (mean; 95% CI) | reference | 9,071 (8,671 to 9,470) | 14,095 (13,679 to 14,511) |
| Incremental QALYs, whole-population direct (consistency check) | reference | 8,389 | 15,171 |
| Incremental cost (CEA, off-ART cost ratio 0·45), ¥ billion, base case | reference | -54·4 | -82·5 |
| Incremental cost, PSA (mean; 95% CrI), ¥ billion | reference | -54·9 (-73·8 to -38·7) | -83·3 (-111·6 to -59·3) |
| Incremental QALYs, PSA (mean; 95% CrI) | reference | 8,462 (6,345 to 11,086) | 15,303 (11,769 to 19,823) |
| Net monetary benefit at ¥5 million/QALY, ¥ billion, base case | reference | 99·7 | 153·0 |
| ICER | reference | Dominant | Dominant |
| Replicates dominant (paired per-seed costs and QALYs) | reference | 199/200 | 200/200 |
| Replicates with NMB > 0 | reference | 200/200 | 200/200 |
| Probability of dominance, PSA | reference | 100% | 100% |
| **Budget impact (discounted)** |  |  |  |
| Break-even year | reference | 12 | 11 |
| 40-year cumulative budget impact, ¥ billion (off-ART cost ratio 30%) | reference | -48·7 | -74·8 |

### Uncertainty and convergence

Two distinct sources of uncertainty were addressed. First-order (stochastic, Monte Carlo) uncertainty was characterised by running the model across 200 random-number seeds; incremental outcomes are reported as the mean across seeds with 95% confidence intervals (normal approximation), and the dominance result as the fraction of seeds in which immediate ART was dominant. At 200 seeds the relative Monte Carlo standard error of every principal estimand was approximately 3%, and running-estimate convergence is shown in Supplementary Material section 5. Second-order uncertainty in the health-economic parameters (unit costs, off-ART cost ratio, and health-state utilities) was characterised by probabilistic sensitivity analysis (PSA), one-way deterministic sensitivity analysis (DSA, tornado), and a cost-effectiveness acceptability curve (CEAC), computed on the seed-averaged inputs (Supplementary Material section 6). The transmission, testing, and behavioural parameters were fixed at their calibrated values and were not varied in the probabilistic analysis. Structural uncertainty was explored by varying the analytic-period force-of-infection multiplier across five settings while holding the calibrated 2023 burn-in state fixed, which brackets a range of plausible future epidemic intensities (Supplementary Material section 9).

### Software, reproducibility, and ethics

The microsimulation was implemented in Python 3.11.15 with Numba 0.64.0 just-in-time acceleration (model file hiv_rapid_art_model.py); analyses used NumPy 2.4.3, pandas 3.0.1, SciPy 1.17.1, Matplotlib 3.10.8, and seaborn 0.13.2, and were run with fixed, reported random-number seeds. Model code and selected aggregated outputs are openly available (https://github.com/toshtanig/Rapid_ART_ABM) and archived on Zenodo (https://doi.org/10.5281/zenodo.21931762). The NDB claims analysis that produced the aggregated cost inputs was approved by the Institutional Review Board of Chiba University (approval number M10805), and only publication-cleared aggregate NDB outputs were used in this study. The simulation itself used publicly available aggregate surveillance data and published estimates, and no new individual-level data were collected for this study.

## Results

The calibrated model reproduced the contemporary Japanese epidemic at the end of the burn-in period (2023). Modelled annual HIV and AIDS notifications were 911 per year (95% range across seeds 823 to 1,000; observed approximately 950 for 2022 to 2024), the number on ART was 29,931 (29,042 to 30,820; NDB and core-hospital estimates approximately 26,000 to 30,000), the undiagnosed fraction was 11·2% (approximately 10 to 12; estimated national range 10 to 20%), viral suppression among those on ART was approximately 99·7% (reported above 99%), and the heterosexual share of incident infections was approximately 19% (approximately 16% in surveillance) (Supplementary Figure S1; Table 1; Supplementary Material section 2). The calibration also reproduced the historical rise and fall of national notifications, including the 2008 to 2013 peak (modelled peak approximately 1,570 around 2013 against an observed peak of approximately 1,590), providing a face-validity check of the calibrated transmission trajectory; because the historical trajectory was itself a calibration target, this is a time-series fit rather than independent hold-out validation. The time-varying burn-in calibration is described in full, with year-by-year factors, in Supplementary Material section 2.

Under the one-month delay, the modelled diagnosis-to-ART interval had a median of 30 days (IQR 30 to 61), consistent with the observed median of about 42 days,^15^ which fell within the modelled interquartile range. Its mean was longer (120 days, 3·9 months) because of a right-tailed subgroup diagnosed at CD4 of 500 or above with HIV-RNA below 5,000 copies/mL, who must await clinical progression to qualify for certification. The eligibility-to-ART interval under the one-month delay, governed by the four-week statutory requirement, had a median of 30 days (IQR 30 to 61) and a mean of 48 days (1·6 months). The immediate and two-month strategies produced diagnosis-to-ART medians of 0 days (mean about 1 day) and 66 days (IQR 61 to 91; mean 154 days), respectively (Table 1; Supplementary Figure S6).

Over the 40-year horizon the cumulative number of new HIV infections rose from 13,263 under immediate ART to 16,254 under the one-month delay (2,991 additional infections) and 17,797 under the two-month delay (4,535 additional infections). Cumulative deaths among people with HIV showed a smaller absolute gradient (32,576, 34,337, and 35,263 respectively), and cumulative ART initiations rose with delay (25,662, 27,843, and 29,030) because the larger pool of incident infections in delayed strategies eventually fed back into the diagnosis-and-ART stream (Table 1). Annual new infections remained consistently lower under immediate ART throughout the projection, with widening separation in cumulative infections through the horizon (Figure 2).

**Figure 2.**
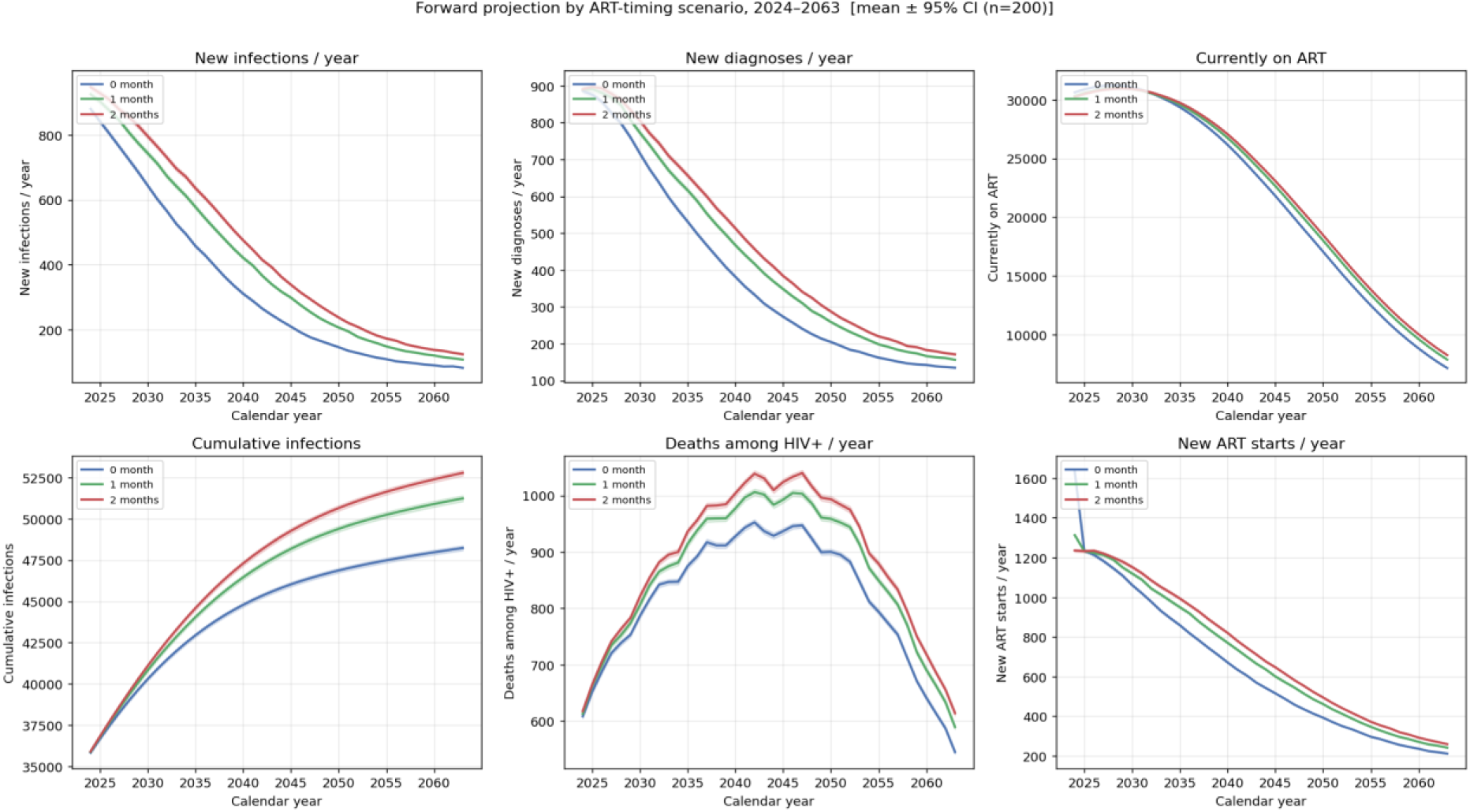
Projected annual epidemic outcomes by ART-initiation strategy, 2024 to 2063. Mean (line) with 95% confidence interval (band) across 200 seeds for the immediate (0 month), one-month, and two-month strategies: (A) new infections per year, (B) new diagnoses per year, (C) people currently on ART, (D) cumulative infections, (E) deaths among people with HIV per year, and (F) new ART initiations per year.

Immediate ART was dominant at the base case; across 200 replicates, the net monetary benefit was positive in all 200, and paired per-seed incremental costs and QALYs showed dominance in 199 of 200 under either cost convention (Table 1; Figure 3). Immediate ART averted a mean of 2,991 cumulative HIV infections (95% CI 2,871 to 3,112) and 1,761 deaths among people with HIV (95% CI 1,686 to 1,837) over 40 years. The base-case incremental QALY gain, estimated by the prespecified prevention decomposition, was 9,071 (95% CI 8,671 to 9,470), comprising a prevention component of 8,406 and a treatment-timing component of 665; the whole-population direct estimate was concordant at 8,389 (the anchor of the decomposition’s survival weight). Immediate ART reduced cumulative discounted healthcare costs by ¥54·4 billion (the whole-population difference in total discounted costs), and the net monetary benefit at ¥5 million per QALY was ¥99·7 billion. The discounted loss per averted infection was approximately 2·8 QALYs. In probabilistic sensitivity analysis, the mean cost saving was ¥54·9 billion (95% credible interval 38·7 to 73·8 billion) and the mean QALY gain 8,462 (6,345 to 11,086), with dominance in all 1,000 iterations (Supplementary Figure S4; Table 1).

**Figure 3.**
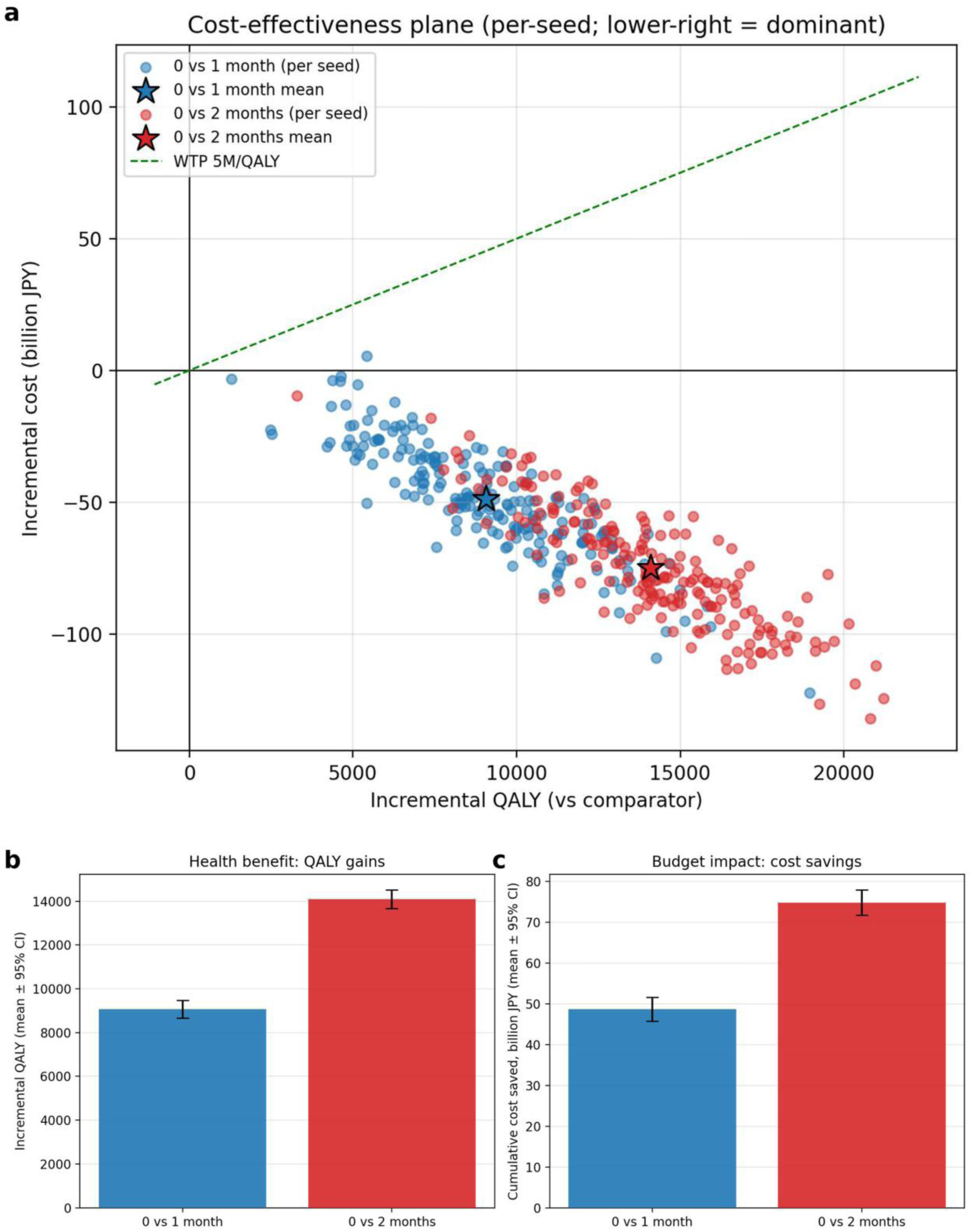
Incremental cost-effectiveness of immediate ART. Panel A, cost-effectiveness plane showing each seed as a point (incremental QALYs versus incremental cost) for immediate ART versus the one-month and two-month delays, with the mean and the willingness-to-pay line; points in the lower-right quadrant indicate dominance. Panel B, health benefit (incremental QALYs) and Panel C, budget impact (cumulative cost savings), each as the mean with 95% confidence interval by comparison. Incremental costs in Panels A and C are paired per-seed differences computed under the budget-impact cost convention (off-ART cost ratio 0·30); the CEA base case (off-ART cost ratio 0·45) and the probabilistic results, in which costs and the off-ART ratio are sampled from distributions centred on the base-case values, are reported in Table 1 and Supplementary Figure S4.

Compared with the two-month delay, which together with the one-month delay brackets the observed interval of about six weeks, immediate ART was dominant in 200 of 200 replicates and cost-saving in all 200. Immediate ART averted a mean of 4,535 cumulative infections (95% CI 4,411 to 4,658) and 2,688 deaths (95% CI 2,610 to 2,766), gained 14,095 QALYs (95% CI 13,679 to 14,511; whole-population direct estimate 15,171), and reduced cumulative discounted healthcare costs by ¥82·5 billion (whole-population total-cost difference; probabilistic sensitivity analysis mean ¥83·3 billion, 95% credible interval 59·3 to 111·6 billion), with a net monetary benefit of ¥153·0 billion. A pairwise comparison of the one-month against the two-month delay also showed dominance of the shorter delay (direct estimate 6,782 additional QALYs and ¥28·2 billion in cost savings; dominance in all PSA iterations), consistent with a graded benefit as the delay shortens.

At the base case, all pairwise comparisons of earlier versus later ART initiation were dominant, and the cost-effectiveness acceptability curve indicated cost-effectiveness of immediate ART in all iterations at every threshold examined (0 to ¥20 million per QALY; Supplementary Figure S5). In the long-term discounted expenditure projection, a modest initial increase in expenditure was followed by savings from averted infections; cumulative savings offset the early investment at year 12 against the one-month delay and at year 11 against the two-month delay, reaching ¥48·7 billion and ¥74·8 billion by year 40 (Figure 4). In the short-term budget impact, the undiscounted annual additional expenditure in years 1 to 5 was ¥0·7, ¥0·7, ¥0·6, ¥0·5, and ¥0·3 billion against the one-month delay (cumulative ¥2·7 billion) and ¥0·8, ¥0·8, ¥0·7, ¥0·6, and ¥0·4 billion against the two-month delay (cumulative ¥3·3 billion). In one-way deterministic sensitivity analysis, the net monetary benefit was most sensitive to the discount rates for costs and for QALYs and to the off-ART cost ratio, but remained positive across all one-way variations: for the primary comparison the net monetary benefit ranged from ¥76·5 billion to ¥130·3 billion, and for the secondary comparison from ¥128·6 billion to ¥209·4 billion, and no single-parameter perturbation reversed dominance (Figure 4; Supplementary Material section 6).

**Figure 4.**
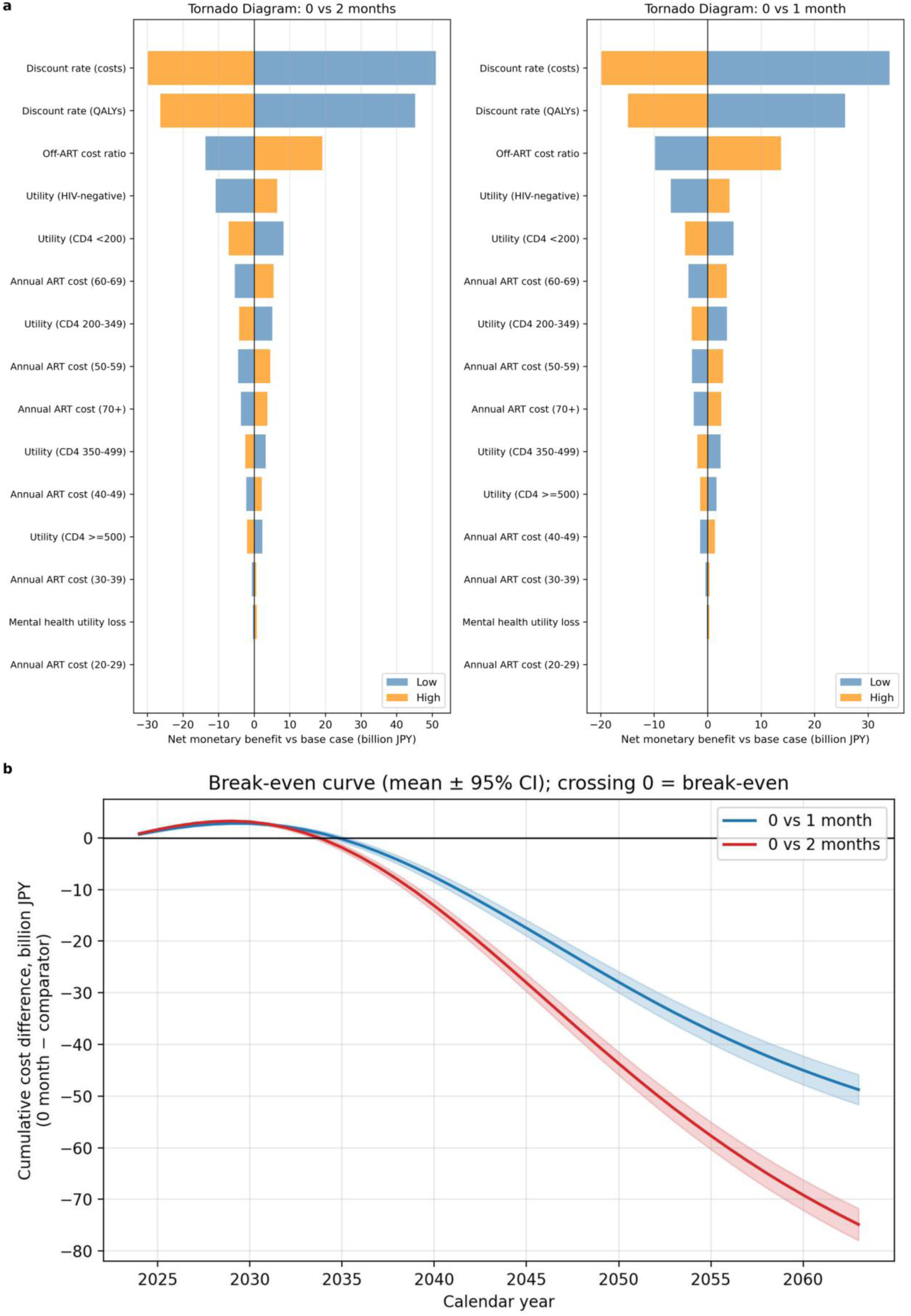
Parameter sensitivity and budget impact. Panel a, one-way deterministic sensitivity analysis (tornado diagrams) of the net monetary benefit of immediate ART versus the two-month delay (left) and the one-month delay (right); the net monetary benefit remained positive across the full range of every parameter (¥76·5 billion to ¥130·3 billion for the primary comparison). Panel b, break-even curve: the cumulative discounted cost difference (immediate minus delayed) over the 40-year horizon, with the year of break-even where the curve crosses zero (year 12 versus the one-month delay; year 11 versus the two-month delay); line, mean; band, 95% confidence interval across 200 seeds. The cost-effectiveness acceptability curve is shown in Supplementary Figure S5.

At the point-estimate level, dominance was retained under both QALY estimators; replicate-level dominance used the prespecified prevention-decomposition estimator, and within the plausible range it did not depend on the counterfactual survival weight. The sign of the QALY gain and of the cost saving was positive in essentially all replicates, and the assumption- independent outcomes (infections averted and cost savings) were tightly estimated (relative Monte Carlo standard error approximately 3%). In a one-way sensitivity analysis varying the counterfactual survival weight across the plausible range (per-infection discounted QALY of approximately 2·2 to 3·5), the incremental QALY scaled approximately linearly while the incremental cost was unchanged, so immediate ART remained dominant with a positive net monetary benefit throughout this range; only the magnitude of the health gain, and not the cost-effectiveness conclusion, was sensitive to this assumption, and dominance attenuated only under an implausible lower-bound stress test in which the per-infection QALY approached zero (Supplementary Material section 4).

In a structural sensitivity analysis (100 seeds) that varied the analytic-period force-of-infection multiplier across five settings from 1·00 to 1·30 while holding the calibrated 2023 burn-in state fixed, immediate ART remained dominant at every setting (in 99 to 100 of 100 replicates); infections averted against the one-month delay ranged from 1,499 (multiplier 1·00) to 5,205 (multiplier 1·30), with 3,072 in the base case (multiplier 1·18), confirming that the absolute effect scales with assumed future epidemic intensity whereas the cost-saving, dominant conclusion was retained across the tested multiplier range (Supplementary Material section 9).

In a 100-seed model-based decomposition of domestic transmission pressure (an attribution of the monthly force of infection by infectious-source state, not a reconstruction of observed transmission chains), the contribution from diagnosed individuals not yet on ART increased from 0·1% under immediate ART to 7·1% under the one-month delay and 9·3% under the two-month delay. Of the additional infections generated by the one-month delay, about 37% were attributed directly to the diagnosed-but-untreated window and about 65% to onward transmission in the still-undiagnosed pool, with the small remainder reflecting a reduction in transmission from individuals transiently on ART before suppression, partly offset by transmission from those disengaged from care (Supplementary Material section 8).

## Discussion

In this microsimulation, calibrated to contemporary Japanese surveillance and cascade estimates, immediate ART, achievable by relaxing or reforming the disability-certification pathway so that all diagnosed individuals can start ART at diagnosis, was projected to dominate the current standard of care on both health and economic grounds over a 40-year horizon. Against the one-month delay, immediate ART prevented 2,991 new infections and 1,761 deaths among people with HIV, gained 9,071 discounted QALYs, and reduced discounted healthcare costs by ¥54·4 billion, and was dominant at the base case, with a positive net monetary benefit in all 200 replicates and dominance in all probabilistic sensitivity-analysis iterations. The secondary comparison against the two-month delay produced a larger absolute benefit (4,535 infections and 2,688 deaths averted; 14,095 discounted QALYs; ¥82·5 billion in savings). Cumulative savings offset the early investment within the second decade (year 12 and year 11). Probabilistic and one-way sensitivity analyses preserved dominance; across the plausible range of the counterfactual survival weight only the magnitude of the health gain changed, and dominance failed only under an extreme lower-bound stress test (Supplementary Material). The model attributed the additional infections under delayed ART to the diagnosed-but-untreated window directly and to amplified onward transmission in the still-undiagnosed pool (Supplementary Material section 8), consistent with part of the benefit being mediated by elimination of this transmission window. Consistent with the concentrated epidemic, the averted infections and deaths were predominantly among MSM (about 87 to 90% of the totals; Supplementary Table S6). Even in a low-prevalence concentrated epidemic with an already strong downstream cascade, shortening the time from diagnosis to ART may deliver clinically meaningful, population-level, and fiscally favourable benefits.

The direction of our findings is consistent with the international evidence base. Randomised trials in sub-Saharan Africa have shown that same-day or same-week ART shortens time to viral suppression and improves engagement in care,^6,7^ and the San Francisco RAPID programme reduced the median time from diagnosis to viral suppression from 134 days to 61 days.^8,9^ A systematic review concluded that rapid ART is associated with shorter time to viral suppression and higher retention without increased early adverse events.^10^ The benefit of earlier treatment for the individual is supported by the START and TEMPRANO trials.^28,29^ Our study extends this literature in two ways. First, we quantify the long-term epidemiological, clinical, and economic benefits of rapid ART in a low-prevalence, concentrated-epidemic, high-income setting with an already high downstream cascade,^12,13^ a context that differs substantially from the settings of most prior evidence. Second, we position the intervention against the specific structural feature that drives the current Japanese diagnosis-to-ART interval, namely the disability-certification pathway. To our knowledge this is the first integrated individual-level evaluation that explicitly represents this pathway and estimates the consequences of its relaxation across epidemiological, clinical, and budget-impact dimensions simultaneously; previous Japanese modelling was compartmental and focused on combination prevention.^16^

The agent-based design integrates transmission, disease progression, testing, the statutory certification pathway, ART timing, retention, PrEP, and mortality within a single individual-level simulation, and represents the two-assessment, four-week certification requirement as an eligibility check at diagnosis followed by a scenario-specific minimum delay with monthly re-evaluation. The one-month and two-month comparators were benchmarked against observed clinical practice and chosen to bracket the observed diagnosis-to-ART interval,^15^ the economic evaluation used NDB-derived treatment costs and EQ-5D-5L population norms,^12,25^ and uncertainty was characterised at two levels, namely stochastic uncertainty across 200 replicates with Monte Carlo standard errors approximately 3% and parameter uncertainty through probabilistic and one-way sensitivity analyses. The model reproduced the heterosexual share of incident infections (approximately 19% versus approximately 16% in surveillance) through an explicit high-activity heterosexual core, addressing a structural difficulty in earlier versions of the model.

Several limitations should be acknowledged. The estimated benefit is the joint effect of removing the CD4/HIV-RNA eligibility criteria and the waiting interval; these components were not decomposed in separate scenarios, although the subgroup affected only by the eligibility criteria is quantified in Supplementary Figure S7 and remains small. First, the size of the high-risk heterosexual at-risk population in Japan is among the most uncertain inputs, and although the concentrated high-activity core reproduces the observed heterosexual share, the absolute heterosexual burden remains uncertain; any underestimate would, if anything, reinforce the dominance of immediate ART by under-counting averted heterosexual infections. Second, the model uses a closed set of behavioural groups with a stationary contact structure, and the 40-year horizon carries projection uncertainty that widens with time; the transmission, testing, and behavioural parameters were fixed at their calibrated values and not propagated in the probabilistic sensitivity analysis, so the dominance result is conditional on the calibrated epidemiological structure; the direction of the effect was nevertheless robust across the tested range of future epidemic intensities, although its absolute magnitude was not (Supplementary Material section 9). Third, reproducing the historical rise and fall of national notifications, including the 2008 to 2013 peak, required a time-varying transmission and testing calibration during the burn-in (Supplementary Material section 2); although each component is anchored to documented testing volumes and to the historical treatment-eligibility thresholds, these year-specific factors add model flexibility; we therefore relied on the incremental design, in which all strategies share the common calibrated burn-in, so that the comparison is not confounded by the initial epidemic state. Fourth, validation of the diagnosis-to-ART interval relied principally on data from a major Designated HIV/AIDS Core Hospital,^15^ with regional variation approximated by the two-month scenario; the observed 42-day median was bracketed by the one-month and two-month delays, across which the conclusion was consistent and graded. Fifth, the payer perspective excludes productivity losses, likely underestimating the total benefit. Healthcare costs were derived from the publication-cleared NDB aggregate for fiscal years 2013 to 2020 and were not adjusted to a common price year, which could alter the magnitude of the projected savings; common-price-year conversion was not examined directly, although the net monetary benefit remained positive across wide cost variation in the deterministic sensitivity analysis.

Antiretroviral prices may fall through generic substitution or pricing revisions, shrinking the projected savings. Utilities for people with HIV were drawn from international cohorts, as no Japanese CD4-stratified utility data were identified; these values were varied widely in the probabilistic and one-way sensitivity analyses. Sixth, we assumed immediate ART can be delivered without additional adverse effects or adherence decrements, consistent with implementation experience,^10^ but local implementation would require attention to counselling, linkage pathways, and psychosocial support. The mortality outcome counts all-cause deaths among people with HIV; part of the reduction reflects fewer people entering the HIV-positive state, and it should not be read as an equivalent reduction in all-cause population mortality.

We assumed no change in sexual behaviour after diagnosis; because the transmission benefit is mediated by viral suppression, this is unlikely to bias the direction of the result. Finally, HIV drug resistance is not represented explicitly; given contemporary integrase-inhibitor-based first-line regimens,^30^ this is unlikely to alter the population-level conclusions.

The disability-certification pathway, although it has served an important historical role in making ART financially accessible under universal health insurance, now functions as an important structural contributor to the diagnosis-to-ART interval, consistent with the observed median delay of about 42 days; in that study, 1·6% of previously untreated patients did not meet the certification criteria and remained untreated, whereas 19·4% of those already receiving ART at first presentation had not obtained the certificate.^15^ Statutory relaxation or reform of the certification pathway to permit immediate ART was projected to prevent approximately 3,000 to 4,500 new HIV infections and 1,760 to 2,690 deaths among people with HIV over 40 years, and to reduce discounted public-payer expenditure by approximately ¥54 to ¥83 billion, consistent across the prespecified sensitivity analyses. Break-even within the second decade indicates that the short-term investment was projected to be recovered well within the analytic horizon. Because Japan has not implemented a rapid ART policy aligned with the WHO recommendation,^5,15^ statutory reform would align national practice with the contemporary international standard, and provides a quantitative evidence base for ongoing policy dialogue on reforming the certification pathway for HIV.

## Contributors

Toshibumi Taniguchi conceived the study, designed the simulation model, developed the model code, conducted the analyses, prepared the figures, drafted the manuscript, and is the guarantor of the work. Mayumi Imahashi contributed clinical input on HIV care delivery and the certification pathway in Japanese practice, contributed to interpretation, and critically revised the manuscript. Tatsuya Noda extracted and analysed the National Database of Health Insurance Claims data underlying the cost parameters, assessed the validity of these data, contributed to interpretation of the health-economic findings, and critically revised the manuscript. Daisuke Sato, as a specialist in health economic evaluation, provided expert assessment and confirmation of the validity of the health-economic evaluation in this study, contributed to interpretation of the findings, and edited the manuscript. Toshibumi Taniguchi and Tatsuya Noda directly accessed and verified the underlying National Database of Health Insurance Claims data. All authors had full access to all the data in the study, contributed to the interpretation of findings, approved the final version, and accept responsibility for the decision to submit for publication.

## Data sharing statement

The model source code (hiv_rapid_art_model.py and the post-processing scripts), the parameter values embedded in the source code and tabulated in Supplementary Tables S1 and S5a, the computational environment specification, and selected aggregated outputs supporting the principal tables and figures are available to anyone, without restriction. The Overview, Design concepts, and Details (ODD) model protocol and the principal parameter tables are provided in the Supplementary Material; further parameter values are documented in the source code. These materials are deposited in a public repository (https://github.com/toshtanig/Rapid_ART_ABM) and are archived with a permanent digital object identifier issued through Zenodo (DOI 10.5281/zenodo.21931762), released under an open licence (the MIT License for code and the Creative Commons Attribution licence [CC BY] for data). No new individual participant data were collected for this study. The individual-level NDB claims records underlying the aggregated cost inputs cannot be shared under the NDB data-provision framework; the aggregated cost inputs themselves are provided in Supplementary Table S1. All calibration targets derive from publicly available aggregate surveillance reports and published care-cascade estimates, and therefore no participant-level data sharing applies. Requests for access before publication may be directed to the corresponding author.

## Data Availability

https://doi.org/10.5281/zenodo.21931762

## Acknowledgments and funding

This study was supported by the Health and Labour Sciences Research Grant, AIDS Control Policy Research Program, Ministry of Health, Labour and Welfare of Japan (grant numbers 21HB1003, 23HB1001, and 26HB1001). The funder had no role in study design; in the collection, analysis, and interpretation of data; in the writing of the report; or in the decision to submit the paper for publication. The authors thank the AIDS Surveillance Committee of Japan for the surveillance work that underpins the calibration of this model, and colleagues at the Designated HIV/AIDS Core Hospitals for clinical and operational insights informing the certification-pathway component of the model.

## Reporting standards

This analysis is reported in accordance with the Consolidated Health Economic Evaluation Reporting Standards 2022 (CHEERS 2022),^18^ the ISPOR-SMDM Modeling Good Research Practices for dynamic transmission modelling,^20^ and the Overview, Design concepts, and Details (ODD) protocol for agent-based models.^19^ The full ODD description appears in Supplementary Material section 1.

