## Supplementary Material for "Treating earlier, spending less: cost-effectiveness and budget impact of immediate versus delayed antiretroviral therapy for HIV in Japan"

This Supplementary Material describes the simulation model following the Overview, Design concepts, and Details (ODD) protocol (section 1), and provides calibration, health-economic, uncertainty, subgroup, transmission-source decomposition, and force-of-infection sensitivity details (sections 2 to 9), with supplementary tables and figure legends. Decimal points are shown midline per journal style.

### Contents


1. Model description following the ODD protocol (Tables S1, S1a, S1b, S1c)

2. Calibration details (Tables S2, S3; Figure S1)

3. Budget impact and break-even analysis

4. Health-economic methods detail (Table S4; Figure S2)

5. Stochastic uncertainty and convergence (Table S5; Figure S3)

6. Parameter uncertainty (Table S5a; Figures S4, S5)

7. Heterogeneity and subgroup analyses (Table S6; Figures S6, S7)

8. Transmission-source decomposition (Tables S7, S8)

9. Analytic-period force-of-infection multiplier sensitivity (Table S9)

Supplementary Figures S1 to S7


### 1. Model description following the ODD protocol

#### 1.1 Purpose

The model is a stochastic individual-based (agent-based) dynamic transmission microsimulation of HIV acquisition, diagnosis, and care in Japan, linked to a cost-effectiveness analysis from the healthcare payer perspective. Its purpose is to estimate the 40-year health, epidemiological, and economic consequences of permitting immediate antiretroviral therapy (ART) by relaxing the statutory immune-dysfunction disability certification pathway, compared with one-month (primary) and two-month (secondary) delays.

#### 1.2 Entities, state variables, and scales

The entities are individual people in two sexual risk populations: men who have sex with men (MSM) and a high-risk heterosexual population (heterosexual men and women) that contains a small high-activity core representing concentrated heterosexual transmission. Each individual carries state variables including sex, age, risk group, high-activity-core membership, foreign-born status, HIV status, CD4 category, plasma HIV-RNA (viral load, log10 copies/mL), diagnosis status, ART status and time on ART, viral suppression, PrEP status, and, for the analytic period, infection and diagnosis cohort-year tags and accumulated discounted QALYs. The spatial scale is national (Japan, unstructured mixing within risk groups); the temporal scale is a monthly time step over a 20-year burn-in (calendar 2004 to 2023) followed by a 40-year analytic horizon (2024 to 2063).

#### 1.3 Process overview and scheduling

Each month, the following processes are executed: demographic turnover (entry of new susceptibles and exogenous foreign-born people with HIV; ageing; background mortality and the excess mortality of people with HIV); PrEP uptake and discontinuation; HIV transmission (a per-capita force of infection by risk group, conditioned on the viral-load distribution of unsuppressed infectious individuals and on per-contact transmission probabilities); disease progression (CD4 decline and viral-load assignment when untreated); HIV testing and diagnosis subject to a national testing-capacity constraint; certification-eligibility evaluation, where applicable, and scheduling of ART initiation with the scenario-specific delay; ART effects (viral suppression and CD4 recovery); and pre-ART and on-ART dropout. Incident-infection and incident-diagnosis cohorts and their discounted QALYs are tracked during the analytic period only. To minimise Monte Carlo variation between scenarios, event-specific independent random-number streams are re-seeded identically across scenarios (a common-random-numbers approximation), so that between-scenario differences are attributable solely to ART timing.

#### 1.4 Design concepts

Basic principles: treatment as prevention, whereby viral suppression reduces onward transmission, links individual ART timing to population-level incidence. Emergence: the epidemic trajectory and the heterosexual share of incidence emerge from individual transmission events rather than being imposed. Stochasticity: testing, infection, progression, mortality, and behavioural transitions are random processes; results are pooled across 200 random-number seeds. Observation: monthly and yearly aggregates (new infections, diagnoses, deaths, people on ART, undiagnosed fraction) and incident-cohort QALYs are recorded. The model has no adaptive learning or explicit networks beyond risk-group and high-activity-core structure.

#### 1.5 Initialisation

The burn-in starts from an estimated early-2000s state with 16,500 prevalent people with HIV, of whom 7,500 are on ART, allocated across MSM, heterosexual men, and heterosexual women in the ratio 0·75 / 0·125 / 0·125. Susceptible populations are initialised at 1,000,000 MSM and 1,000,000 high-risk heterosexual individuals (target 2,100,000 with catch-up entry). The high-risk heterosexual population of 1,000,000 is a scenario assumption representing a broad high-risk heterosexual exposure population (for example commercial-sex clients, sex workers, sexually transmitted infection clinic attendees, and people with multiple partners) rather than a directly measured official estimate, for which none is available in Japan; its order of magnitude is indirectly consistent with Japanese behavioural and sex-industry data. Because the heterosexual share of incidence (about 16 to 19%) is reproduced by calibrating the contact rate and the high-activity core, the between-strategy contrasts are insensitive to this absolute size. The 20-year burn-in brings the simulated epidemic to a contemporary state calibrated to 2023; the burn-in end-state is then forked identically into the three ART-timing scenarios.

#### 1.6 Input data

Calibration targets and demographic, behavioural, and cost inputs are drawn from publicly available aggregate sources: the AIDS Surveillance Committee of Japan annual report (2024); National Database of Health Insurance Claims (NDB) care-cascade estimates; published per-act transmission meta-analyses; CD4 natural-history studies; treatment-era cohort standardised mortality ratios; Japanese EQ-5D-5L population norms and a CD4-stratified utility compendium; and the MHLW AIDS Control Policy Research project for the diagnosis-to-ART interval and NDB-derived costs. No individual participant data are used. Full values and sources are listed in Table S1.

#### 1.7 Submodels

Force of infection. For each risk group, the monthly per-susceptible hazard is proportional to the group contact rate and to the prevalence of unsuppressed infectious partners weighted by viral-load-stratified per-act transmission probabilities. Viral-load bins are defined on the log10 scale with edges 0, 2·3, 4·0, 5·0, 6·0, and 10·0. Per-act probabilities are 0, 0·001, 0·001, 0·002, and 0·005 for MSM and 0, 0·00015, 0·0003, 0·0006, and 0·0015 for heterosexual contacts (upper end of published confidence intervals). The high-activity heterosexual core (3% of the heterosexual population) has a contact-rate multiplier of 14, an initial HIV enrichment weight of 12, and a foreign-born share of 0·15, reproducing concentrated bridge transmission. Individuals with treatment-induced viral suppression below the modelled suppression threshold (HIV-RNA at or below 1·5 log10 copies/mL, approximately 30 copies/mL) contributed effectively no transmission and were excluded as infectious sources, consistent with undetectable equals untransmittable; untreated individuals with low but detectable viral load were assigned the per-act transmission probability of the corresponding viral-load bin shown above. A phenomenological calibration multiplier was applied to the force of infection (1·18 during the analytic period and year-specific during the burn-in); to compensate for the reduced infectious pool introduced by the CD4- and care-state-stratified mortality, model_144 additionally applies uniform recalibration scalars to the force of infection (a factor of 1·05 throughout and an additional factor of 1·04 from 2016 onward), so that the analytic-period multiplier of 1·18 corresponds to an effective analytic-period force of infection of approximately 1·29; the year-by-year burn-in factors are given in Supplementary Material section 2.

Testing, diagnosis, certification, and ART initiation. Monthly testing probabilities depend on HIV status, CD4 category, risk group, age, and PrEP status, subject to a national testing-capacity constraint that scales from 14,500 to 18,000 tests per month over five years in the analytic period; the lower value was set to approximate recent national screening volumes and the upper value the planned national expansion target, and during the burn-in the testing capacity followed documented national administrative HIV-testing volumes (Supplementary

Material section 2). This analytic-period testing-capacity assumption is a modelling choice; because diagnosis precedes the ART-timing contrast identically in all strategies, the between-strategy comparison is not driven by it, although it affects the absolute projected epidemic size. In the immediate-ART scenario, all diagnosed individuals initiated ART in the month of diagnosis irrespective of CD4 count or HIV-RNA; neither certification eligibility nor the associated waiting interval was applied. In the one-month and two-month comparator scenarios, the statutory certification criteria (CD4 below 500 cells/microlitre, or CD4 of 500 or above with HIV-RNA of 5000 copies/mL or above) were applied. Individuals meeting the criteria at diagnosis were assigned a time to ART drawn from a distribution with mean equal to the scenario target (1 or 2 months) and standard deviation 0·5 months (the eligibility-to-ART interval), whereas individuals not meeting the criteria remained in monthly follow-up until they became eligible once CD4 declined below 500 or HIV-RNA rose to 5000 or above; the diagnosis-to-ART interval therefore additionally includes this waiting time.

CD4 and viral-load dynamics. Untreated individuals progress through five CD4 stages with stochastic decline and viral load assigned by CD4 category; after ART initiation, viral load declines to suppression and CD4 recovers, with monthly viral-load variation modelled during treatment.

PrEP. Oral PrEP uptake grows over ten years to a maximum of 2·0% among MSM (from 1·0%) and 0·5% among heterosexual individuals (from 0%), with 95% effectiveness and a monthly discontinuation probability of 0·5%.

Mortality and demographic turnover. Each living person with HIV is assigned, in every monthly cycle, an age- and sex-specific general-population background mortality hazard multiplied by a relative risk (a standardised mortality ratio, SMR) determined jointly by the current CD4 stratum and care state; the monthly death probability is p = 1 minus exp(minus background hazard times relative risk). The full CD4- and care-state-stratified schedule, its sources, and an internal validation are given in section 1·8 (Tables S1a to S1c). New susceptibles enter at an annualised 2·0% with a catch-up mechanism to the target population, and foreign-born people with HIV enter as an exogenous monthly process with separate burn-in and analytic-period rates (burn-in 25 / 30 / 50 and analytic 38 / 16 / 13 per year for MSM, heterosexual men, and heterosexual women).

Pre-ART and on-ART dropout are applied at monthly probabilities of 0·01 and 0·001, respectively.

**Table S1. Model parameters, values, and sources**

| **Parameter** | **Value** | **Source / note** |
|---|---|---|
| MSM population (burn-in start) | 1,000,000 | Network scale-up estimates of MSM in Japan[1] |
| High-risk heterosexual population | 1,000,000 | Scenario assumption (no official estimate); broad high-risk heterosexual exposure population. Heterosexual incidence share reproduced by calibration, not by this absolute size (section 1·5)[2,3,4] |
| Target population (catch-up) | 2,100,000 | Demographic catch-up target |
| New-entrant rate | 2·0% per year | Demographic turnover assumption |
| Burn-in / analytic horizon | 20 years / 40 years | 2004–2023 burn-in; 2024–2063 analysis |
| Initial prevalent HIV / on ART | 16,500 / 7,500 | Estimated early-2000s state (AIDS Surveillance cumulative reports)[2] |
| Initial HIV allocation (MSM/HetM/HetF) | 0·75 / 0·125 / 0·125 | Concentrated MSM epidemic with heterosexual bridge |
| Contact rate (MSM / heterosexual) | 10·1 / 7·0 | Calibrated to 2020–2024 notifications and MSM:heterosexual ratio[2] |
| VL bins (log10 edges) | 0, 2·3, 4·0, 5·0, 6·0, 10·0 | Viral-load stratification |
| Per-act probability, MSM | 0, 0·001, 0·001, 0·002, 0·005 | Patel 2014; PARTNER and Opposites Attract[5–7] |
| Per-act probability, heterosexual | 0, 0·00015, 0·0003, 0·0006, 0·0015 | Boily 2009 (upper CI)[8]; calibrated bridge transmission |
| High-activity core: share / activity / HIV weight / foreign share | 3% / ×14 / ×12 / 0·15 | Concentrated heterosexual bridge calibration |
| Testing capacity, analytic period (per month, over 5 years) | 14,500 → 18,000 | Model assumption (planned national testing expansion); burn-in capacity anchored to documented administrative testing volumes, year-specific (section 2) |
| Transmission calibration multiplier | 1·18 (analytic); year-specific in burn-in | Phenomenological calibration of the force of infection reproducing the national notification trajectory; with uniform recalibration scalars (×1·05 throughout, ×1·04 from 2016) the effective analytic-period force of infection is approximately 1·29 (section 2) |

| Parameter | Value | Source / note |
|---|---|---|
| All-cause mortality among people with HIV (RR/SMR by CD4 stratum and care state) | see Table S1a | CD4- and care-state-stratified standardised mortality ratios; ART-CC[20]; Lodwick[21]; COHERE[22]; Japanese validity check Konishi[23] (section 1·8) |
| Burn-in CD4 ART-initiation threshold | ≤350 (2004–2008); ≤500 (2009–2013); none thereafter | Historical treatment-eligibility transition (section 2) |
| Foreign-born HIV influx (burn-in; analytic), MSM/HetM/HetF per year | 25/30/50; 38/16/13 | AIDS Surveillance Committee 2024 imported-transmission estimates[2] |
| ART-timing scenarios (mean, SD months) | 0 (0); 1 (0·5); 2 (0·5) | Eligibility-to-ART delay by scenario |
| Pre-ART / on-ART monthly dropout | 0·01 / 0·001 | Japanese retention data[9,10] |
| PrEP max uptake MSM / heterosexual | 2·0% / 0·5% over 10 years | Japanese PrEP context[11]; iPrEx effectiveness 95%[12]; discontinuation 0·5%/month |
| Discount rate (costs and QALYs) | 2·0% per year | Japanese (C2H) guidance[13] |
| HIV-negative utility (U_NEG) | 0·94 (prevention decomposition) | Representative HIV-negative utility for the prevention-decomposition counterfactual; the cost-effectiveness analysis instead uses age-dependent HIV-negative utilities (about 0·86 to 0·95). Shiroiwa 2021 EQ-5D-5L norms[14] |
| Utilities by CD4 (≥500/350–499/200–349/<200) | 0·87 / 0·83 / 0·76 / 0·65 | Poku 2025 compendium[15]; Japanese value set; international cohorts (no Japanese CD4-stratified utility data identified) |
| Mental-health utility decrement during the diagnosed pre-ART period | 0·06, applied pro rata by months | Shiroiwa 2021[14]; Brandt 2017[16] |
| Annual on-ART cost (age-stratified) | ¥2·4 to ¥3·4 million by age | NDB FY2013–2020, publication-cleared aggregate[9,17] |
| Off-ART cost ratio | 0·45 (0·30 in budget-impact analysis) | Reduced cost reflecting absence of ART drug cost; antiretroviral medication is the dominant HIV care cost component, and Japanese inpatient/outpatient HIV cost data[18,19] |
| Counterfactual survival weight (SURVIVAL_ADJ) | 0·648 | Anchored to whole-population direct ΔQALY (section 4) |
| Willingness-to-pay threshold | ¥5,000,000 per QALY | Base-case WTP |
| Replicates (random-number seeds) | 200 | Common random numbers across scenarios |

#### 1.8 CD4- and care-state-stratified all-cause mortality among people with HIV

In every monthly cycle each living person with HIV is assigned an age- and sex-specific general-population background mortality hazard that is multiplied by a relative risk determined jointly by the current CD4 stratum and care state. The monthly death probability is p = 1 minus exp(minus background hazard times relative risk). The relative-risk values are standardised mortality ratios (SMRs) relative to the age- and sex-matched general population; because an SMR is an all-cause mortality ratio, it is conceptually consistent with applying the multiplier to the background hazard. Four CD4 strata (500 or above, 350 to 499, 200 to 349, and below 200 cells/microlitre) and four care states (untreated, on ART with viral suppression, on ART without viral suppression, and disengaged from care) define a 4 by 4 schedule (Table S1a). Because the Japanese HIV epidemic is concentrated among MSM, MSM-specific SMRs were used wherever available.

**Table S1a. All-cause mortality relative risk among people with HIV (standardised mortality ratio relative to the general population), by care state and CD4 stratum**

| Care state | CD4 ≥500 | CD4 350–499 | CD4 200–349 | CD4 <200 |
|---|---|---|---|---|
| Untreated (no ART) | 1·0 | 1·4 | 5·0* | 10·0* |
| On ART, virally suppressed | 1·0 | 1·1 | 1·5 | 2·6 |
| On ART, not suppressed | 2·0 | 2·1 | 3·5 | 5·5 |
| Disengaged from care | 1·5* | 2·0* | 6·0* | 12·0* |

*Extrapolated value (assumption flag): no directly observed CD4-and-care-state-specific stratified SMR suitable for this cell was identified; the value was set to preserve a monotonic CD4 gradient and natural-history severity, and is a designated target of sensitivity analysis (Supplementary Material section 4 and section 9). The two directly anchored cells are the suppressed and non-suppressed CD4 200 to 349 values (1·5 and 3·5, from ART-CC). The high-CD4 suppressed and non-suppressed cells are splits of the directly reported ART-CC CD4 of 350 or above SMRs across the 500-or-above and 350-to-499 strata using the reported high-CD4 gradient; the pooled CD4 below 200 values are person-time-weighted representatives of the finer ART-CC CD4 cells (100 to 199, 50 to 99, 0 to 49), most model person-time being near 100 to 199.

The schedule was anchored to treatment-era cohort SMRs and checked for domestic validity. The suppressed and non-suppressed rows are anchored to the Antiretroviral Therapy Cohort Collaboration (ART-CC), which reported SMRs for MSM (CDC stage A/B) by 6-month CD4 and 6-month viral load relative to the general population of nine industrialised countries (Table S1b).[20] The untreated CD4 of 350 or above values are anchored to Lodwick and colleagues, who reported an SMR of 1·30 (95% CI 1·06 to 1·58) for ART-naive MSM with CD4 above 350, with an adjusted rate ratio of 0·77 for CD4 500 to 699 versus 350 to 499 (implying a near-background value at CD4 500 or above).[21] A residual CD4 gradient under viral suppression is supported by COHERE.[22] Recent Japanese data (men with HIV, 2021 to 2024) provide a domestic validity check: an SMR of about 1·05 at CD4 of 500 or above, about 1·03 at viral load below 50 copies/mL, about 2·59 at CD4 below 200, about 3·37 at viral load 1000 copies/mL or above, and about 10·43 for those with treatment interruption or who were untreated.[23] The untreated CD4 below 350 cells and all disengaged-from-care cells were not directly available as stratified SMRs and were extrapolated, set above the corresponding untreated or non-suppressed values to represent advanced or disengaged disease while preserving monotonicity, and remain principal targets of the structural and survival sensitivity analyses.

**Table S1b. ART-CC source SMRs (MSM, CDC stage A/B), by 6-month CD4 and 6-month viral load**

| 6-month CD4 (cells/microlitre) | Viral load ≤500 (suppressed) | Viral load >500 (not suppressed) |
|---|---|---|
| ≥350 | 1·05 (0·82–1·35) | 2·06 (1·37–3·10) |
| 200–349 | 1·47 (1·07–2·01) | 3·50 (2·33–5·27) |
| 100–199 | 2·27 (1·57–3·29) | 3·52 (2·09–5·95) |
| 50–99 | 3·23 (1·68–6·21) | 6·59 (2·96–14·7) |
| 0–49 | 3·41 (0·85–13·7) | 23·4 (13·0–42·2) |

Source: ART-CC (Int J Epidemiol 2009).[20] Values are SMRs (95% CI) for MSM, CDC clinical stage A or B.

**Table S1c. Internal validation: person-years, mortality rate, and observed-to-expected ratio by care state and CD4 stratum (immediate-ART scenario, 40-year horizon)**

| Care state | CD4 | Person-years | Deaths per 1000 person-years | O/E |
|---|---|---|---|---|
| Untreated | ≥500 | 40,074 | 15·6 | 0·98 |
| Untreated | 350–499 | 37,426 | 18·0 | 0·96 |
| Untreated | 200–349 | 48,096 | 51·0 | 0·98 |
| Untreated | <200 | 58,870 | 111·8 | 1·02 |
| On ART, suppressed | ≥500 | 880,145 | 28·8 | 1·00 |
| On ART, suppressed | 350–499 | 615,928 | 27·2 | 0·99 |
| On ART, suppressed | 200–349 | 643,909 | 31·7 | 1·00 |
| On ART, suppressed | <200 | 402,640 | 43·3 | 1·01 |
| On ART, not suppressed | ≥500 | 250 | 40·1 | 1·28 |
| On ART, not suppressed | 350–499 | 455 | 15·4 | 0·59 |
| On ART, not suppressed | 200–349 | 953 | 34·6 | 1·05 |
| On ART, not suppressed | <200 | 3,071 | 62·5 | 1·12 |
| Disengaged from care | ≥500 | 28,221 | 42·0 | 1·02 |
| Disengaged from care | 350–499 | 11,886 | 53·8 | 1·05 |
| Disengaged from care | 200–349 | 7,570 | 130·9 | 1·04 |
| Disengaged from care | <200 | 4,997 | 192·7 | 0·95 |

The observed-to-expected (O/E) ratio was close to 1·0 in 13 of 16 cells (pooled mean 1·00), confirming that the implementation reproduces the specified mortality structure; the three cells with wider deviation are the non-suppressed high-CD4 cells, which accrue very little person-time and are therefore statistically noisy.

### 2. Calibration details

The burn-in was calibrated so that the modelled epidemic at the end of the burn-in (calendar year 2023) reproduced the contemporary Japanese HIV epidemic, and the burn-in trajectory reproduced the historical rise and fall of national notifications. Calibration jointly adjusted contact rates, the heterosexual viral-load-stratified per-act probabilities, the high-activity-core parameters, foreign-born influx, and the initial prevalent allocation, together with a year-specific time-varying calibration of the force of infection and of the burn-in testing capacity, and a historical CD4-count threshold governing ART initiation during the burn-in. The burn-in testing capacity was anchored to documented national administrative HIV-testing volumes, and the historical CD4-count ART-initiation thresholds (CD4 of 350 cells/microlitre or below for 2004 to 2008 and 500 cells/microlitre or below for 2009 to 2013) reflect the treat-all transition; the full year-by-year calibration factors are tabulated below. Calibration was performed by iterative manual adjustment rather than a formal optimisation algorithm: these parameters were tuned so that the burn-in end-state (calendar year 2023) reproduced contemporary national targets within their observed ranges, prioritising annual notifications, the undiagnosed count, and the number on ART, while reproducing the observed national notification trajectory. The calibration targets were annual HIV notifications of about 654, AIDS notifications of about 292, and total notifications of about 946; AIDS at diagnosis of about 31%; transmission-route shares of about 80% for MSM and 20% for heterosexual exposure; about 29,680 people currently on ART (acceptance range 28,500 to 30,500); viral suppression on ART above 99%; and about 3,500 undiagnosed people with HIV (range 3,200 to 3,800), with 33,000 to 36,000 people with HIV overall as a soft target. A parameter set was accepted when the seed-averaged 2023 values fell within these ranges. As a face-validity check, the model reproduced the observed historical rise to the 2008 to 2013 notification peak (modelled peak approximately 1,570 around 2013 against an observed peak of approximately 1,590) and the subsequent decline. Achieved values at 2023 (mean across 200 seeds; 95% CI) are shown in Table S2, and the year-by-year fit against national data in Supplementary Figure S1. The time-varying transmission, testing-capacity, and ART-eligibility calibration factors are a phenomenological device. Because all strategies share the same calibrated burn-in, the incremental comparison is not confounded by differences in the initial epidemic state; however, the absolute magnitude of infections averted and cost savings depends on the calibrated future epidemic intensity, as explored in Supplementary Material section 9.

**Table S2. Calibration targets and modelled values at 2023**

| Indicator | Modelled (95% range across seeds) | Observed / target |
|---|---|---|
| Annual HIV and AIDS notifications | 911 (823 to 1,000) | approximately 950 (2022–2024) |
| People currently on ART | 29,931 (29,042 to 30,820) | approximately 26,000 to 30,000 (NDB, core hospitals) |
| Undiagnosed fraction | 11·2% (approximately 10 to 12) | 10 to 20% (national estimate) |
| Viral suppression on ART | approximately 99·7% | greater than 99% |
| Heterosexual share of incident infections | approximately 19% | approximately 16% (surveillance) |

**Table S3. Year-by-year burn-in calibration factors (2004 to 2023)**

| Year | FOI multiplier | Monthly testing capacity | Undiagnosed testing boost | ART CD4 threshold (cells/microlitre) | Modelled notifications |
|---|---|---|---|---|---|
| 2004 | 1·00 | 14,500 | 1·00 | ≤350 | 1,030 |
| 2005 | 1·00 | 15,000 | 1·00 | ≤350 | 1,148 |
| 2006 | 1·00 | 15,500 | 1·00 | ≤350 | 1,204 |
| 2007 | 1·00 | 16,000 | 1·00 | ≤350 | 1,247 |
| 2008 | 1·00 | 16,500 | 1·00 | ≤350 | 1,278 |
| 2009 | 1·02 | 16,500 | 1·00 | ≤500 | 1,285 |
| 2010 | 1·04 | 17,500 | 1·05 | ≤500 | 1,360 |
| 2011 | 1·08 | 18,500 | 1·10 | ≤500 | 1,415 |
| 2012 | 1·12 | 19,500 | 1·20 | ≤500 | 1,490 |
| 2013 | 1·15 | 20,500 | 1·35 | ≤500 | 1,572 |
| 2014 | 1·12 | 20,000 | 1·20 | none (universal) | 1,355 |
| 2015 | 1·09 | 18,500 | 1·10 | none (universal) | 1,190 |
| 2016 | 1·07 | 18,000 | 1·05 | none (universal) | 1,111 |
| 2017 | 1·07 | 17,500 | 1·00 | none (universal) | 1,051 |
| 2018 | 1·07 | 17,000 | 1·00 | none (universal) | 1,026 |
| 2019 | 1·07 | 17,000 | 1·00 | none (universal) | 1,007 |
| 2020 | 1·06 | 14,500 | 1·00 | none (universal) | 948 |
| 2021 | 1·06 | 14,000 | 1·00 | none (universal) | 918 |
| 2022 | 1·07 | 14,500 | 1·00 | none (universal) | 922 |
| 2023 | 1·07 | 15,000 | 1·00 | none (universal) | 911 |

FOI, force of infection. The tabulated force-of-infection multiplier is the year-specific burn-in calibration curve; the analytic period (2024 onward) used a constant multiplier of 1·18, which with the uniform recalibration scalars (1·05 throughout and 1·04 from 2016 onward) corresponds to an effective analytic-period force of infection of approximately 1·29, and a testing capacity scaling from 14,500 to 18,000 tests per month. The undiagnosed testing boost is the year-specific multiplier applied to the testing probability of undiagnosed HIV-positive individuals (1·00 outside 2010 to 2016). The ART CD4 threshold is the historical eligibility rule governing burn-in ART initiation (universal thereafter). Modelled notifications are the seed-averaged annual HIV and AIDS notifications; the observed national series is shown against the modelled values in Supplementary Figure S1. The burn-in testing capacities are anchored to documented national administrative HIV-testing volumes.

### 3. Budget impact and break-even analysis

The budget-impact analysis applied the payer perspective with off-ART costs at 30% of on-ART costs to disregard the diagnosis-to-ART transient cost surge. In the short-term budget impact (undiscounted annual cash flows), the additional expenditure in years 1 to 5 was ¥0·7, ¥0·7, ¥0·6, ¥0·5, and ¥0·3 billion against the one-month delay (cumulative ¥2·7 billion) and ¥0·8, ¥0·8, ¥0·7, ¥0·6, and ¥0·4 billion against the two-month delay (cumulative ¥3·3 billion). In the long-term discounted expenditure projection (2% per year), a modest initial increase in expenditure was followed by savings from averted infections; cumulative discounted savings offset the early investment at year 12 against the one-month delay and at year 11 against the two-month delay, within the modelled payer-cost framework and excluding implementation costs, reaching ¥48·7 billion and ¥74·8 billion by year 40 (Figure 4, panel b).

### 4. Health-economic methods detail

Whole-population QALYs. Because the intervention changes who becomes infected, QALYs were accrued over the whole simulated population to capture the benefit of averted infections; restricting QALYs to people with HIV would omit this prevention benefit and can reverse the sign of the estimate.

Utility provenance. Japanese EQ-5D-5L population norms were used for HIV-negative person-time. The CD4-stratified utilities for people with HIV were derived from international cohorts (the Poku 2025 compendium, supplemented by trial-based estimates; supplementary reference 15), as no Japanese CD4-stratified utility data were identified. The dependence of results on these values was examined in the probabilistic and one-way sensitivity analyses (Table S5a).

Prevention decomposition. To obtain a low-variance estimate with confidence intervals, incremental QALYs were also computed as averted infections multiplied by the discounted QALY lost per infection (the difference between counterfactual HIV-negative QALYs and observed post-infection QALYs of the incident-infection cohort), plus a treatment-timing component among individuals infected in both strategies. This estimator targets the same whole-population contrast as the direct difference; with the counterfactual survival weight anchored to the whole-population direct estimate, it provides a low-variance estimate because unaffected individuals contribute zero.

Counterfactual survival weight (SURVIVAL_ADJ). The counterfactual HIV-negative survival weighting was set to 0·648 by anchoring the prevention-decomposition ΔQALY to the whole-population direct estimate (which does not depend on this weight), giving a discounted loss of approximately 2·8 QALYs per averted infection, consistent with near-normal survival under modern ART. A one-way sensitivity analysis (Table S4) varied this weight. SURVIVAL_ADJ scales only the QALY side; incremental cost is unchanged. Across the plausible range (SURVIVAL_ADJ about 0·55 to 0·70) immediate ART remained dominant with a positive net monetary benefit. At the lower-bound stress test (0·45) the dominance fraction falls (173/200), but the incremental QALY remains positive on average and the net monetary benefit remains positive because the cost saving is unaffected (Supplementary Figure S2).

**Table S4. One-way sensitivity to the counterfactual survival weight (0 month vs 1 month; N = 200)**

| SURVIVAL_ADJ | Per-infection QALY | ΔQALY (95% CI) | NMB ¥ billion (95% CI) | Dominant (CEA incremental cost held fixed) |
|---|---|---|---|---|
| 0·45 | 0·3 | 906 (784 to 1,028) | 58·9 (58·3 to 59·5) | 173/200 |
| 0·55 | 1·5 | 5,030 (4,794 to 5,265) | 79·5 (78·3 to 80·7) | 200/200 |
| 0·648 (base) | 2·8 | 9,071 (8,671 to 9,470) | 99·7 (97·7 to 101·7) | 200/200 |
| 0·70 | 3·6 | 11,215 (10,724 to 11,705) | 110·5 (108·0 to 112·9) | 200/200 |
| 0·85 | 5·6 | 17,400 (16,642 to 18,158) | 141·4 (137·6 to 145·2) | 200/200 |

Net monetary benefit is the willingness-to-pay (¥5,000,000 per QALY) multiplied by the incremental QALYs (which vary with SURVIVAL_ADJ) plus the cost-effectiveness incremental cost saving of ¥54·4 billion (whole-population difference, off-ART cost ratio 0·45), which is independent of SURVIVAL_ADJ; the base-case value therefore matches the main-text net monetary benefit (¥99·7 billion). Confidence intervals reflect the across-seed variation in incremental QALYs. The base-case value (0·648) was anchored to the whole-population direct incremental-QALY estimate. Across the plausible range (per-infection discounted QALY of approximately 2 to 3, corresponding to SURVIVAL_ADJ of about 0·55 to 0·70), immediate ART remained dominant and the net monetary benefit remained positive throughout; only the magnitude of the health gain was sensitive to this assumption. The value SURVIVAL_ADJ = 0·45 is a lower-bound stress test (per-infection QALY approaching zero), at which the incremental QALY remains positive on average and the net monetary benefit remains positive although the dominance fraction falls; it is shown to demonstrate robustness rather than as a plausible value.

### 5. Stochastic uncertainty and convergence

Stochastic (Monte Carlo) uncertainty was characterised across 200 random-number seeds; the running mean with 95% CI and the running dominance rate are shown in Supplementary Figure S3. At 200 seeds the relative Monte Carlo standard error of every principal estimand was at most 2·9% (Table S5); under the budget-impact cost convention (0·30) the corresponding maxima were similar (at most 3·1%).

**Table S5. Relative Monte Carlo standard error at 200 seeds (incremental cost and NMB under the CEA cost convention, off-ART ratio 0·45)**

| Comparison | Infections averted | Incremental QALY | Incremental cost | NMB |
|---|---|---|---|---|
| 0 month vs 1 month | 2·05% | 2·25% | 2·85% | 2·50% |
| 0 month vs 2 months | 1·39% | 1·51% | 1·99% | 1·71% |

### 6. Parameter uncertainty

Parameter uncertainty was characterised by probabilistic sensitivity analysis (PSA), one-way deterministic sensitivity analysis (DSA, tornado), and a cost-effectiveness acceptability curve (CEAC), computed with the established cost-effectiveness engine applied to seed-averaged inputs. In PSA, immediate ART was dominant in all 1,000 iterations for both comparisons, with mean cost savings of ¥54·9 billion (primary) and ¥83·3 billion (secondary). The PSA used 1000 Monte Carlo iterations, sampling annual costs from gamma distributions, the off-ART cost ratio and health-state utilities from beta distributions, and the discount rates from normal distributions, with base values, standard errors, distributions, and one-way ranges as listed in Table S5a. The CEAC showed cost-effectiveness in all 1,000 iterations at every willingness-to-pay threshold from 0 to ¥20 million per QALY. In one-way DSA, the net monetary benefit was most sensitive to the discount rates for costs and QALYs and to the off-ART cost ratio, but remained positive across all variations (¥76·5 billion to ¥130·3 billion for the primary comparison; ¥128·6 billion to ¥209·4 billion for the secondary), with no single-parameter perturbation reversing dominance. The probabilistic sensitivity analysis is shown in Supplementary Figure S4 and the cost-effectiveness acceptability curve in Supplementary Figure S5; the full one-way tornado appears as Figure 4 (panel a) in the main text.

**Table S5a. Health-economic parameters in the probabilistic (PSA) and one-way deterministic (DSA) sensitivity analyses**

| Parameter | Base value | SE | Distribution | One-way range |
|---|---|---|---|---|
| Annual on-ART cost, age 20–29 (¥) | 2,375,000 | 475,000 | Gamma | ±20% |
| Annual on-ART cost, age 30–39 (¥) | 2,901,449 | 580,290 | Gamma | ±20% |
| Annual on-ART cost, age 40–49 (¥) | 3,264,767 | 652,953 | Gamma | ±20% |
| Annual on-ART cost, age 50–59 (¥) | 3,433,598 | 686,720 | Gamma | ±20% |
| Annual on-ART cost, age 60–69 (¥) | 3,356,900 | 671,380 | Gamma | ±20% |
| Annual on-ART cost, age 70+ (¥) | 3,164,654 | 632,931 | Gamma | ±20% |
| Off-ART cost ratio (CEA) | 0·45 | 0·10 | Beta | 0·20–0·80 |
| Utility, HIV-negative (age 30–39 anchor) | 0·950 | 0·02 | Beta | 0·90–0·98 |
| Utility, CD4 ≥500 (suppressed on ART) | 0·87 | 0·03 | Beta | 0·80–0·93 |
| Utility, CD4 350–499 | 0·83 | 0·04 | Beta | 0·74–0·90 |
| Utility, CD4 200–349 | 0·76 | 0·05 | Beta | 0·65–0·85 |
| Utility, CD4 <200 | 0·65 | 0·06 | Beta | 0·50–0·78 |
| Mental-health utility decrement (pre-ART period, applied pro rata by months) | 0·06 | 0·02 | Beta | 0·03–0·12 |
| Discount rate, costs (annual) | 0·02 | 0·005 | Normal | 0·00–0·04 |
| Discount rate, QALYs (annual) | 0·02 | 0·005 | Normal | 0·00–0·04 |

SE, standard error. One-way ranges are the prespecified DSA ranges; for the age-stratified annual on-ART costs, the DSA varied the base value by ±20% (SE, 20% of the base). Annual costs were sampled from gamma, the off-ART cost ratio and utilities from beta, and discount rates from normal distributions. The budget-impact analysis used an off-ART cost ratio of 0·30. Costs are in Japanese yen at the prices of the NDB source period (fiscal years 2013 to 2020). CD4-stratified utilities for people with HIV derive from international cohorts (no Japanese CD4-stratified utility data identified); Japanese population norms are used for HIV-negative person-time.

### 7. Heterogeneity and subgroup analyses

Outcomes were tracked separately for MSM and heterosexual individuals (model option ENABLE_HETEROGENEITY_TRACKING). The benefits of immediate ART in averted infections and in fewer all-cause deaths among people with HIV were concentrated in the MSM population, consistent with the concentrated Japanese epidemic: against the one-month delay, MSM accounted for 2,650 of 2,991 averted infections (about 89%) and 1,584 of 1,761 averted all-cause deaths among people with HIV (about 90%), the remainder occurring among heterosexual individuals (Table S6). The heterosexual contribution was small in absolute terms but reproduced the observed heterosexual share of incidence (approximately 19%). The cumulative distribution of the diagnosis-to-ART and eligibility-to-ART intervals by strategy (Supplementary Figure S6) confirms the modelled delays. Supplementary Figure S7 shows the size of the off-ART subgroup with high CD4 (500 cells/microlitre or above) and low viral load (HIV-RNA below 5000 copies/mL) among individuals diagnosed for at least 24 months. This subgroup has a different composition by strategy. In the one-month and two-month scenarios it consists predominantly of individuals who did not meet the statutory certification criteria and therefore remained untreated under the eligibility rule. In the immediate-ART scenario, where ART is initiated at diagnosis irrespective of CD4 and HIV-RNA, no individual is withheld from treatment by the criteria; the residual subgroup therefore comprises individuals who initiated ART and subsequently disengaged from care (treatment discontinuation), and its smaller size reflects the elimination of the criteria-driven untreated reservoir.

**Table S6. Averted infections and all-cause deaths among people with HIV by risk group (mean across 200 seeds; 95% CI)**

| Risk group | Infections averted, 0m vs 1m | All-cause deaths among people with HIV averted, 0m vs 1m | Infections averted, 0m vs 2m | All-cause deaths among people with HIV averted, 0m vs 2m |
|---|---|---|---|---|
| MSM | 2,650 (2,548 to 2,753) | 1,584 (1,517 to 1,651) | 3,926 (3,816 to 4,037) | 2,369 (2,297 to 2,441) |
| Heterosexual | 341 (290 to 392) | 177 (149 to 205) | 608 (551 to 666) | 319 (287 to 351) |
| Total | 2,991 | 1,761 | 4,535 | 2,688 |

Averted = (delayed strategy) minus (immediate ART), as the paired per-seed difference. About 87 to 90% of averted infections and of averted all-cause deaths among people with HIV were among MSM, consistent with the concentrated epidemic. QALY and cost benefits follow the same concentration because they are driven by averted infections; group-specific QALY and cost were not estimated separately.

### 8. Transmission-source decomposition

To identify the care-state origin of domestic transmission pressure, the monthly force of infection was attributed to the infectious-source state of the transmitting person with HIV, accumulated over the 40-year horizon for each strategy. Because the model does not retain individual transmission pairs, this is a model-based attribution of transmission pressure rather than a reconstruction of observed transmission chains. The four infectious-source states are mutually exclusive and exhaustive: undiagnosed; diagnosed but not yet on ART (the certification-related pre-ART window); on ART but not yet virally suppressed; and disengaged from care after diagnosis. Exogenous foreign-born HIV-positive entries are an external inflow (about 2,677 cumulative entries over 40 years) and are not part of the domestic-transmission denominator. Values are pooled across 100 stochastic replicates.

**Table S7. Cumulative domestic infections over 40 years by transmitting-source state and strategy (mean across seeds; per cent of domestic total in parentheses)**

| Strategy | Undiagnosed | Diagnosed, pre-ART | On ART, not suppressed | Disengaged | Domestic total | Exogenous foreign inflow |
|---|---|---|---|---|---|---|
| Immediate ART (0 month) | 12,667 (95·9%) | 8 (0·1%) | 470 (3·6%) | 58 (0·4%) | 13,203 | 2,677 |
| 1-month delay | 14,660 (90·1%) | 1,149 (7·1%) | 154 (0·9%) | 312 (1·9%) | 16,275 | 2,681 |
| 2-month delay | 15,597 (87·8%) | 1,647 (9·3%) | 162 (0·9%) | 366 (2·1%) | 17,772 | 2,675 |

Across all strategies the great majority of domestic transmission arises from undiagnosed individuals (88 to 96 per cent), underscoring that case-finding remains the dominant lever on incidence. The signature of the certification-related delay is the diagnosed-but-pre-ART source, which contributes only 0·1 per cent of infections under immediate ART but rises to 7·1 per cent under the one-month delay and 9·3 per cent under the two-month delay: the transmission window that immediate ART removes.

**Table S8. Additional cumulative domestic infections under delayed versus immediate ART, decomposed by transmitting-source state (mean across seeds; per cent of the additional total in parentheses)**

| Comparison | Additional domestic infections | Undiagnosed | Diagnosed, pre-ART | On ART, not suppressed | Disengaged |
|---|---|---|---|---|---|
| 1-month delay vs immediate | 3,072 | 1,993 (64·9%) | 1,141 (37·1%) | −316 (−10·3%) | 254 (8·3%) |
| 2-month delay vs immediate | 4,569 | 2,930 (64·1%) | 1,639 (35·9%) | −308 (−6·7%) | 308 (6·7%) |

Of the additional infections generated by the one-month delay, about 37 per cent are attributed directly to the diagnosed-but-untreated window and about 65 per cent to onward transmission in the still-undiagnosed pool that grows as the epidemic is sustained at a higher level. A small negative contribution from the on-ART-not-suppressed state (about minus 10 per cent) reflects that, under delay, fewer people have reached ART and therefore less transmission arises from the transiently unsuppressed on-ART state; this is partly offset by a small positive contribution from those disengaged from care (about plus 8 per cent). After including the negative component the contributions sum to the total additional infections. The two-month delay shows the same pattern with larger absolute numbers.

### 9. Analytic-period force-of-infection multiplier sensitivity

The future epidemic intensity over the 40-year analytic horizon is not directly observed and is one of the most consequential structural assumptions. To test whether the cost-effectiveness conclusion depends on it, the analytic-period force-of-infection multiplier was varied across five settings (1·00, 1·10, 1·18 base case, 1·25, and 1·30) while the calibrated 2023 burn-in state was held fixed (the year-specific burn-in calibration and the 2023 care cascade are unchanged). The same set of 100 random-number seeds was used at every setting (common random numbers), so that differences across settings reflect the assumed forward transmission intensity rather than stochastic noise. The multiplier shown is the analytic-period transmission multiplier; the effective analytic-period force of infection additionally carries the calibration scalars and equals approximately the multiplier times 1·092 (shown in parentheses). The base case is a multiplier of 1·18 (effective approximately 1·29).

**Table S9. Cost-effectiveness of immediate ART across analytic-period force-of-infection multipliers (100 seeds per setting; mean with 95% CI across seeds; incremental costs and net monetary benefit use the CEA cost convention, off-ART ratio 0·45)**

| Multiplier (effective FOI) | Comparison | Infections averted (95% CI) | Incremental QALY | Cost saving (¥ billion) | NMB, ¥5M/QALY (95% CI) | Dominant seeds |
|---|---|---|---|---|---|---|
| 1·00 (≈1·09) | Immediate vs 1-month | 1,499 (1,414–1,585) | 5,348 | 27·9 | 54·6 (50·7–58·5) | 99/100 |
| 1·00 (≈1·09) | Immediate vs 2-month | 2,167 (2,076–2,258) | 7,920 | 40·1 | 79·7 (75·5–84·0) | 100/100 |
| 1·10 (≈1·20) | Immediate vs 1-month | 2,221 (2,090–2,352) | 7,092 | 39·9 | 75·3 (69·8–80·8) | 99/100 |
| 1·10 (≈1·20) | Immediate vs 2-month | 3,161 (3,052–3,270) | 10,444 | 56·7 | 108·9 (103·9–114·0) | 100/100 |
| 1·18 (≈1·29, base) | Immediate vs 1-month | 3,072 (2,893–3,250) | 9,228 | 56·4 | 102·5 (95·4–109·7) | 99/100 |
| 1·18 (≈1·29, base) | Immediate vs 2-month | 4,569 (4,401–4,737) | 14,250 | 83·4 | 154·7 (147·6–161·8) | 100/100 |
| 1·25 (≈1·37) | Immediate vs 1-month | 4,084 (3,888–4,280) | 11,611 | 70·2 | 128·2 (120·5–135·9) | 100/100 |
| 1·25 (≈1·37) | Immediate vs 2-month | 6,233 (5,967–6,499) | 18,113 | 108·8 | 199·4 (189·7–209·0) | 100/100 |
| 1·30 (≈1·42) | Immediate vs 1-month | 5,205 (4,963–5,448) | 14,137 | 89·1 | 159·8 (150·7–168·9) | 100/100 |
| 1·30 (≈1·42) | Immediate vs 2-month | 7,901 (7,659–8,143) | 21,766 | 136·3 | 245·1 (236·4–253·8) | 100/100 |

Across the entire tested range, infections averted, incremental QALYs, cost savings, and net monetary benefit all increased monotonically with the assumed force of infection, because a more intense future epidemic offers more infections for immediate ART to avert. Immediate ART remained dominant (less costly and more effective) at every setting, in 99 to 100 of 100 replicates for the immediate-versus-one-month comparison and in all 100 for the immediate-versus-two-month comparison. The cost-effectiveness conclusion is therefore robust to the assumed future epidemic intensity; only the absolute magnitude of the benefit, not its direction or the dominance result, depends on this assumption. The lowest setting (multiplier 1·00) is the most conservative stress test, and immediate ART remained cost-saving and dominant even there.

### Supplementary figures

**Figure S1.** Model calibration against Japanese national data, 2004 to 2023. Modelled annual outcomes (line, median across 200 seeds; band, 95% prediction interval across seeds) compared with observed national data (points) for (a) annual HIV and AIDS notifications, (b) people currently on ART, (c) modelled annual new infections, and (d) the undiagnosed fraction (with the estimated national range shaded). Observed data from the AIDS Surveillance Committee annual report (2024) and NDB and core-hospital estimates.

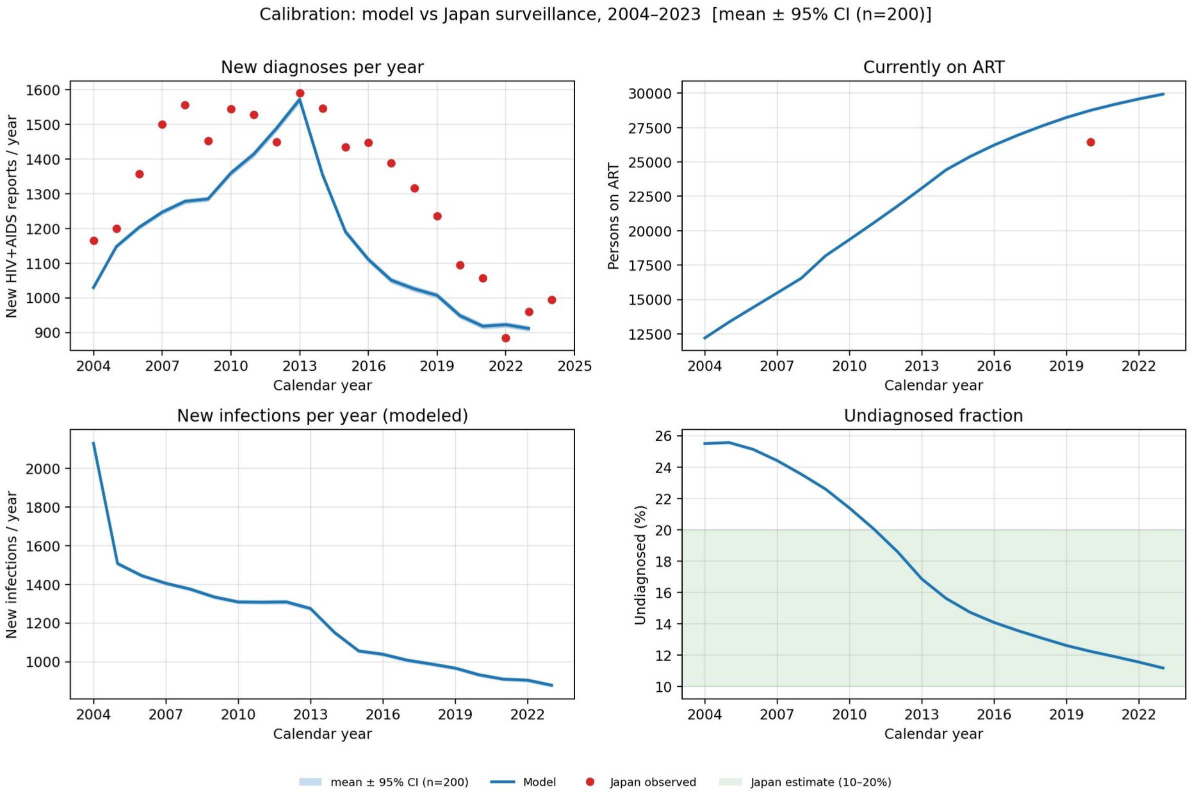

**Figure S2.** One-way sensitivity of per-infection QALYs, incremental QALYs, and net monetary benefit to the counterfactual survival weight (SURVIVAL_ADJ), with the anchored base value (0·648) indicated.

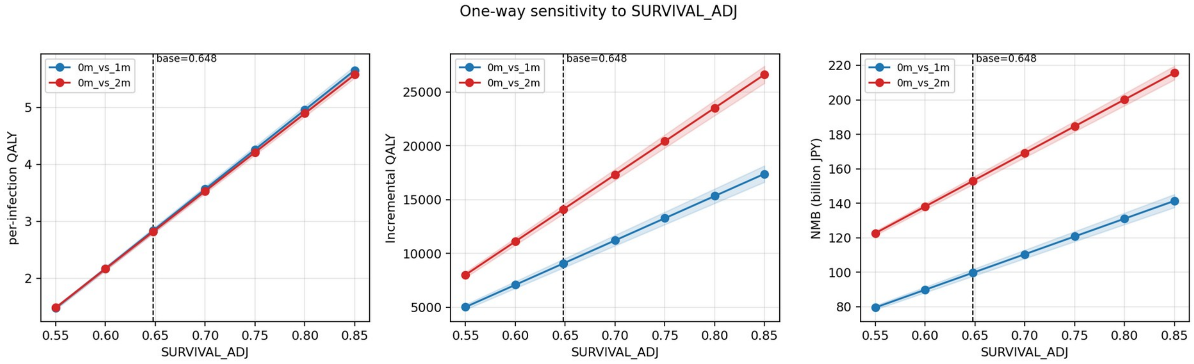

**Figure S3.** Monte Carlo convergence diagnostics: running mean with 95% confidence interval of incremental QALYs and of infections averted, and the running dominance rate, against the number of seeds.

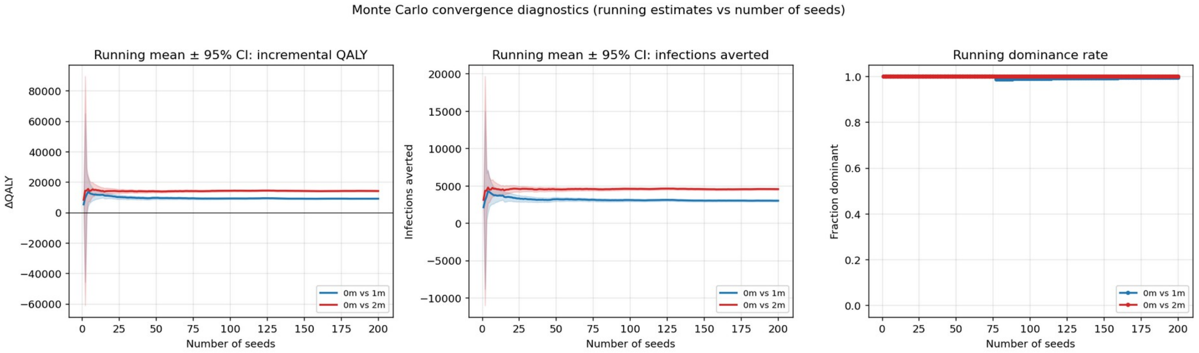

**Figure S4.** Probabilistic sensitivity analysis: (a) cost-effectiveness plane and (b) distributions of incremental cost and incremental QALYs.

a

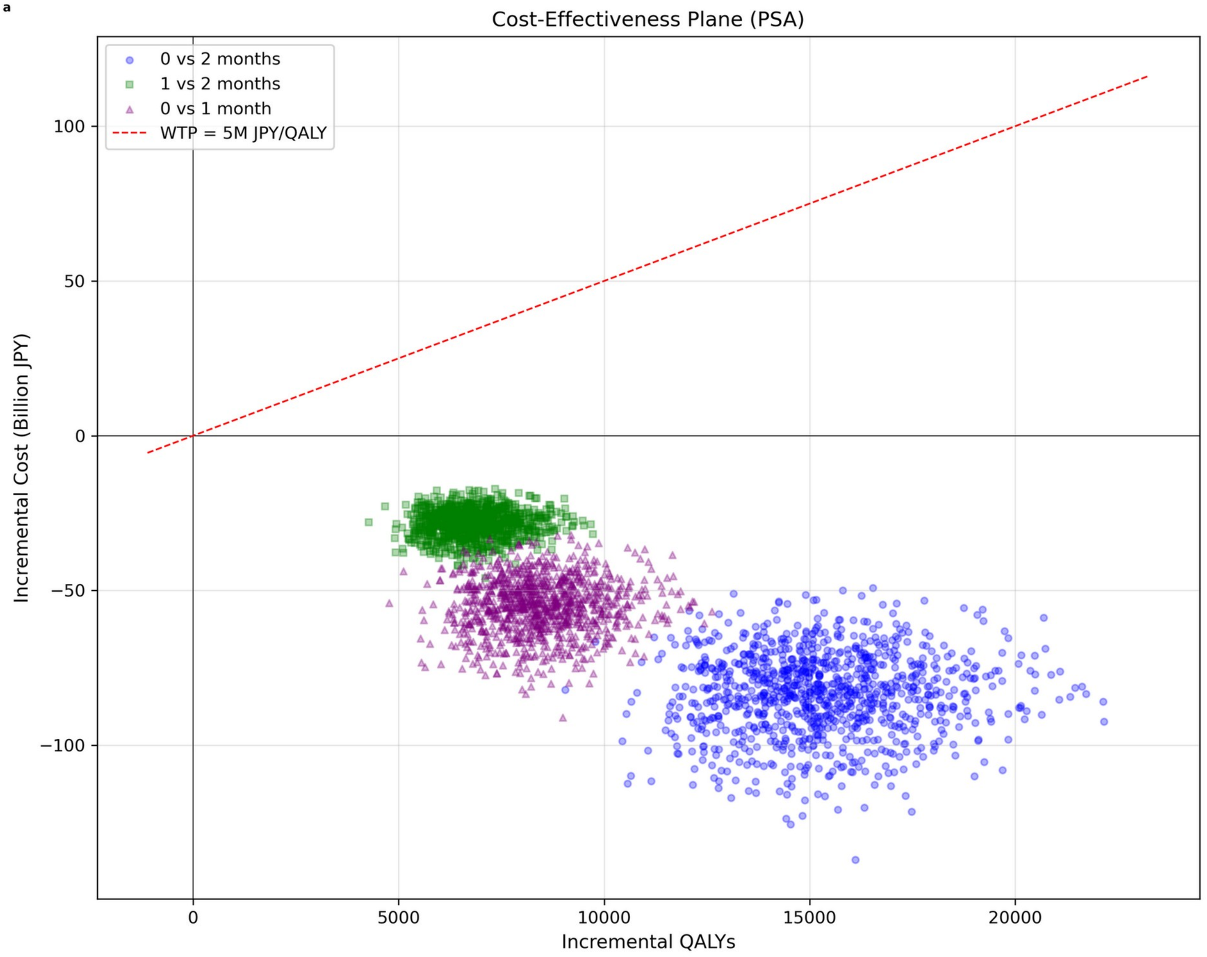


b

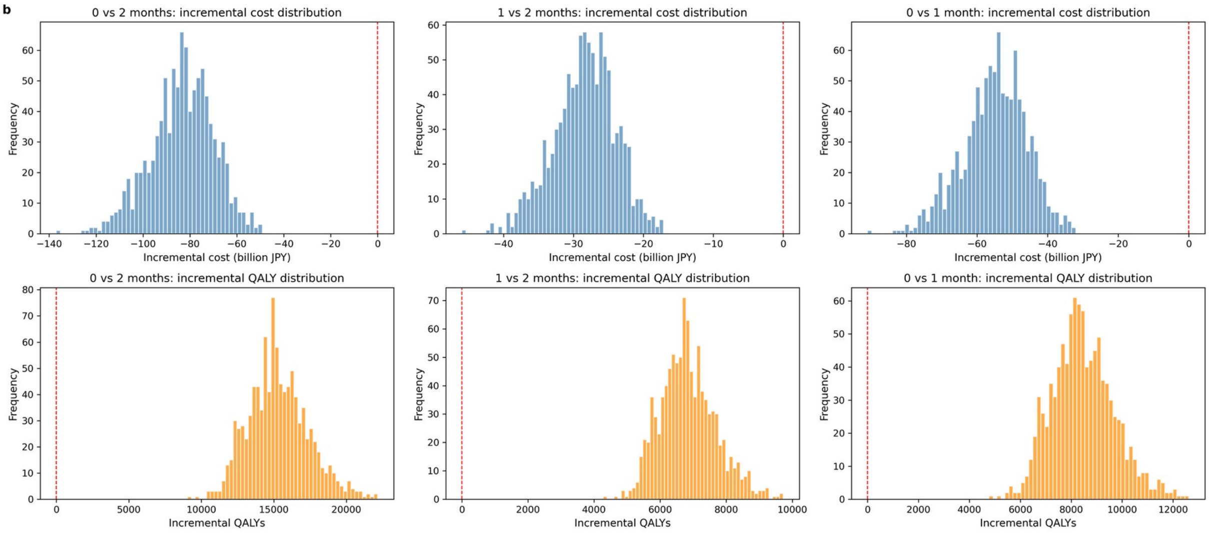

**Figure S5.** Cost-effectiveness acceptability curve across willingness-to-pay thresholds (0 to ¥20 million per QALY): probability that immediate ART is cost-effective for each pairwise comparison.

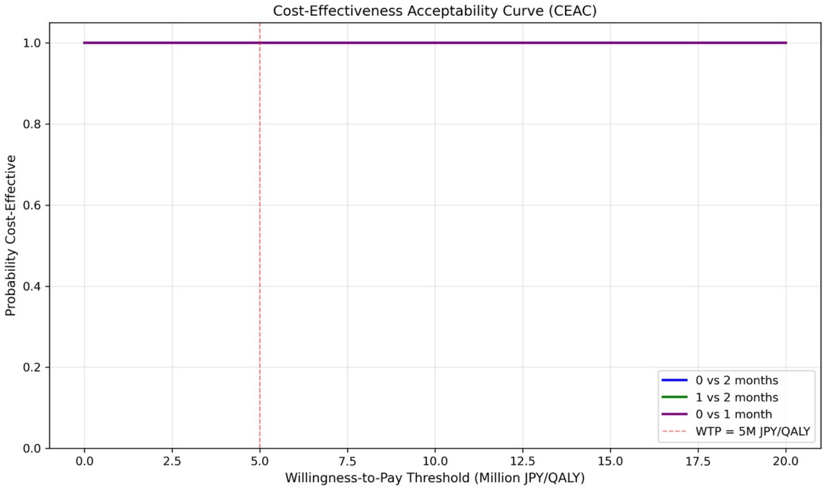

**Figure S6.** Cumulative distribution of (a) the diagnosis-to-ART interval and (b) the eligibility-to-ART interval by strategy (individuals pooled across 200 seeds), confirming the modelled delays.

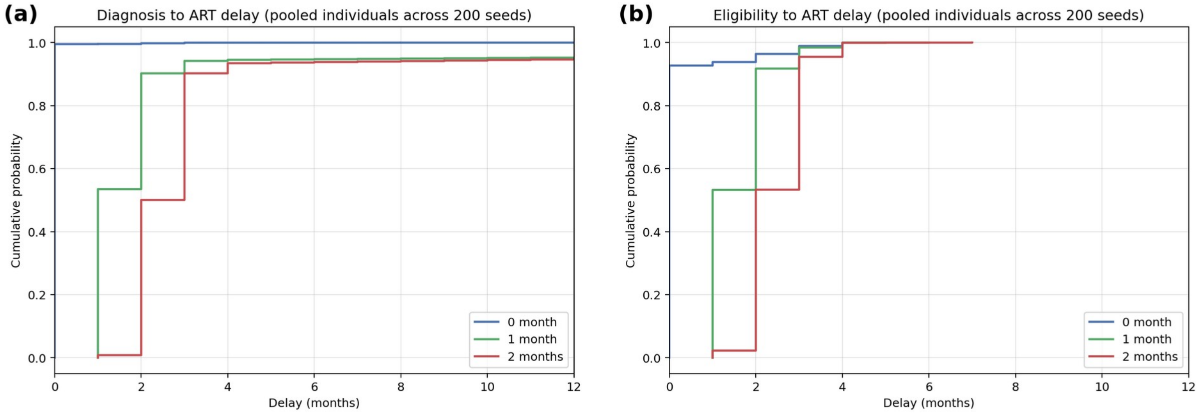

**Figure S7.** Size of the off-ART subgroup with CD4 of 500 cells/microlitre or above and HIV-RNA below 5000 copies/mL among individuals diagnosed for at least 24 months, by strategy (mean with 95% confidence interval across 200 seeds). In the one-month and two-month scenarios this subgroup is composed predominantly of individuals not meeting the statutory certification criteria and therefore not yet treated; in the immediate-ART scenario, where ART begins at diagnosis irrespective of CD4 and HIV-RNA, it comprises individuals who initiated ART and subsequently discontinued treatment rather than individuals withheld from treatment by the criteria.

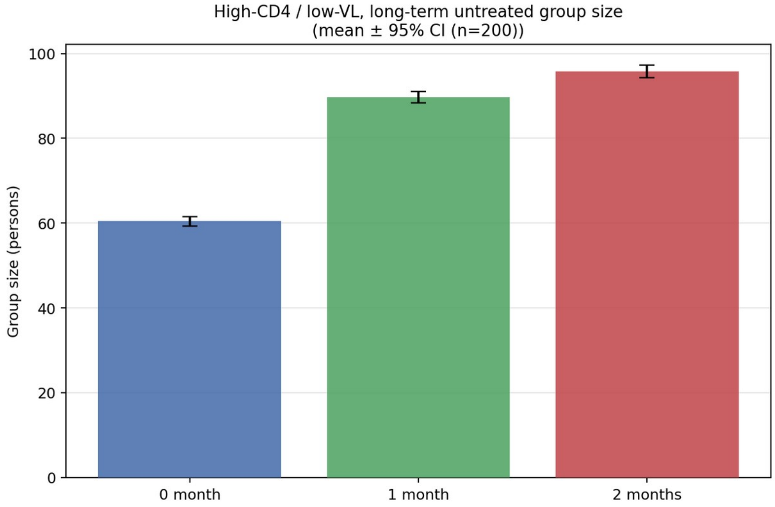
